# Assessing transmission risks and control strategy for monkeypox as an emerging zoonosis in a metropolitan area

**DOI:** 10.1101/2022.06.28.22277038

**Authors:** Pei Yuan, Yi Tan, Liu Yang, Elena Aruffo, Nicholas H. Ogden, Jacques Bélair, Jane Heffernan, Julien Arino, James Watmough, Hélène Carabin, Huaiping Zhu

**Author notes:** These authors contributed equally to this work (Pei Yuan, Yi Tan, Liu Yang). Corresponding Author., 4700 Keele Street, Toronto, Ontario, Canada, M3J1P3.

## Abstract

**Objective:** To model the spread of Monkeypox (MPX) in a metropolitan area for assessing the risk of possible outbreaks, and identifying essential public health measures to contain the virus spread.

**Methods:** The animal reservoir is the key element in the modelling of a zoonotic disease. Using a one health approach, we model the spread of the MPX virus in humans considering animal hosts like rodents (e.g., rats, mice, squirrels, chipmunks, etc.) and emphasize their role and transmission of the virus in a high-risk group, including gay and bisexual men-who-have-sex-with-men (gbMSM). From model and sensitivity analysis, we identify key public health factors and present scenarios under different transmission assumptions.

**Findings:** We find that the MPX virus may spill over from gbMSM high-risk groups to broader populations if efficiency of transmission increases in the higher-risk group. However, the risk of outbreak can be greatly reduced if at least 65% of symptomatic cases can be isolated and their contacts traced and quarantined. In addition, infections in an animal reservoir will exacerbate MPX transmission risk in the human population.

**Conclusion:** Regions or communities with a higher proportion of gbMSM individuals need greater public health attention. Tracing and quarantine (or “effective quarantine” by post-exposure vaccination) of contacts with MPX cases in high-risk groups would have a significant effect on controlling the spreading. Also, surveillance for animal infections would be prudent.

## Introduction

Monkeypox (MPX) is a viral zoonosis first identified in 1958 in a laboratory monkey. Recently, the disease has regained public concern due to the emergence of multiple cases in non-epidemic areas and the prominence of sexual transmission routes^1,2^. At the time of writing, the current outbreak has lasted close to two months since MPX cases were reported in the United Kingdom on May 6, 2022. Since then, there have been an increasing number of non-endemic regions reporting MPX cases including Canada, Spain, Germany, Portugal and the United States. As of June 27, 2022, there have been close to 3413 laboratory confirmed cases from 50 countries/territories in five WHO regions, mainly in non-endemic regions^3^, while the majority of cases from previous outbreaks were found in Central and West Africa^3^. Given the current spreading of the virus, public health agencies of many countries monitor the situation and although the World Health Organisation (WHO) has stated that “the outbreak should not constitute a Public Health Emergency of International Concern (PHEIC) at this stage, however, it acknowledged the emergency nature of the event and that controlling the further spread of this outbreak requires intense response efforts”^5^.

MPX symptoms, similar to those of smallpox, involve two stages from initial prodromal to the rash sub-stage. Initial symptoms are typically flu-like, including fever, chills, exhaustion, headache and muscle weakness^6^. These symptoms are followed by a widespread rash on the face and body, including inside the mouth and on the palms of the hands and soles of the feet^6^. After onset of symptoms, MPX infection can progress to a more severe stage or even death. In outbreaks in Africa, the case fatality rate varies from 1% to 10%^7^.

As an emerging zoonosis, humans can be infected with MPX through close contact with infected humans, animals and the environment^8^. The virus can be transmitted by bodily fluids, such as saliva from coughing (health care workers without appropriate PPE may be vulnerable) and direct physical contact, including sexual contact^8^. Some studies indicated the possibility of transmission from mother to fetus (through close contact during and after birth), but with very limited evidence^8^. In addition, the risk of becoming infected with MPX does not depend only on the mode of transmission, but also on the location where individuals live or work. For example, laboratory personnel and health care workers could have a higher risk^9^, given the high levels of exposure in the workplace. However, the vast majority of cases in the current outbreaks have been reported in the gay, bisexual and other men-who-have-sex-with-men (gbMSM) communities, which was not normally seen in previous outbreaks^10,11,12^. This is quite possibly linked to a virus mutation that made its way into highly interconnected sexual networks within the gbMSM community, with much uncertainty of transmission efficiency based on limited and skewed data^10^. Moreover, the possibility of virus spreading through airborne droplets among the human population is fraught in the global society^8^.

The current outbreaks of MPX are not the first documented outside endemic countries. The first occurred in the United States in 2003, resulting in more than 70 people being infected, some of the cases were infected by pets that were in contact with contaminated rodents imported from Ghana^13^. Animal to human infection may occur by a bite or scratch of animals, bush meat preparation, or direct contact with body fluids or lesion material of infected animals^2,8^. Eating inadequately cooked meat and other infected animal products are also risk factors for infection^2^. While the MPX virus can spread among humans, its transmission from animals to humans also causes concerns, especially if the virus establishes in animals outside the Africa^14^. The increasing pattern of the current spreading raises concern for the formation of animal reservoirs in wildlife which may lead to repeated human outbreaks^14^.

Possible animal hosts of MPX include a range of rodents (e.g., rats, mice, hamsters, gerbils, squirrels, chipmunks, etc.) and non-human primates (e.g., monkeys)^8,13^. Animal-animal transmission may be due to close contact with infected animals or animal tissue, and a range of animal species are thought to be able to infect humans^15^. Even though the first recorded cases in each country documenting MPX infections may not come from animals, the animal reservoir may be a key element to MPX control (or control of any other zoonotic disease) as there is also a probability that human transmission could cause the virus to spill back into animals^16,17^. Besides, due to warming and other environmental changes, populations of animal hosts like rodents may burst in favourable seasons, leading to increased risk of transmission in both humans and animals^18^.

All the new risk factors cause concern about such an emerging threat. MPX requires urgent tackling, as suggested by WHO^19^, and the risk of infection in the naive population needs to be assessed to inform the public, even in the presence of uncertainty. Although infection risk in the general population is considered low^10^, the spill over effect from the gbMSM community or other high-risk groups (HRG) should not be ignored, given the possibility of increasing transmission efficiency due to the virus evolution.

Previous studies have shown that the infectiousness of smallpox patients is much greater after the rash has developed, suggesting that rash-motivated isolation supplies a window for effective control of smallpox^20^. Effective contact tracing was proven effective in the control of smallpox^21^, and is, therefore, expected to be effective in controlling MPX. Contact tracing and ring vaccination are two possible tools for containment of the current outbreak^22^. Nevertheless, regarding the risk for the outbreak to spread to the general population, required actions in the current outbreak are not clear yet.

The increasing spread of MPX in non-endemic communities requires an urgent response, in particular in susceptible metropolitan areas. In this study, we will build dynamical models to mimic the spread of MPX as an emerging zoonosis in some hypothetical metropolitan area, including high- and low-risk human-human transmission, and transmission from animals to humans. These considerations allow for an in-depth study of the transmission mechanisms among and between the animal reservoir and humans, leading to the estimation of the risk of outbreaks under assumptions on different transmission routes and possibility, the identification of model parameters that are particularly influential, and public health measures that may be effective in containing virus spread.

## Methods

### Modelling overview

We constructed a dynamical model for MPX transmission within and between human and animal populations. Based on risk assessments in different population categories exposed to MPX virus^9^, the human population was divided into two groups: the low-risk (LRG, i.e. broader population [subscript 1]) and the HRG (members of the gbMSM community with multiple sexual partners [subscript 2]). The natural reservoir of MPX remains unknown, however, African rodents and non-human primates may harbour the virus and infect people^23^. Wild rodent as an example of animal reservoirs was included, but can comprise any of the mentioned potential reservoir species, as well as new reservoirs if were to emerge. The model followed the SEIR framework extended to include Infectious *p* (prodromal phase) – Infectious *I* (rash phase) – Isolated *Q*_*h*_ (infectious) - Isolated *Q*_*S*_ (susceptible) subpopulations (**Figure 1**). The seasonality of rodent population was included based on the White Footed Mouse *Peromyscus leucopus*^24^. The description of model assumptions, variables and parameters were summarised in Tables 1-4 and details of modelling are in Appendix A.

**Figure 1.**
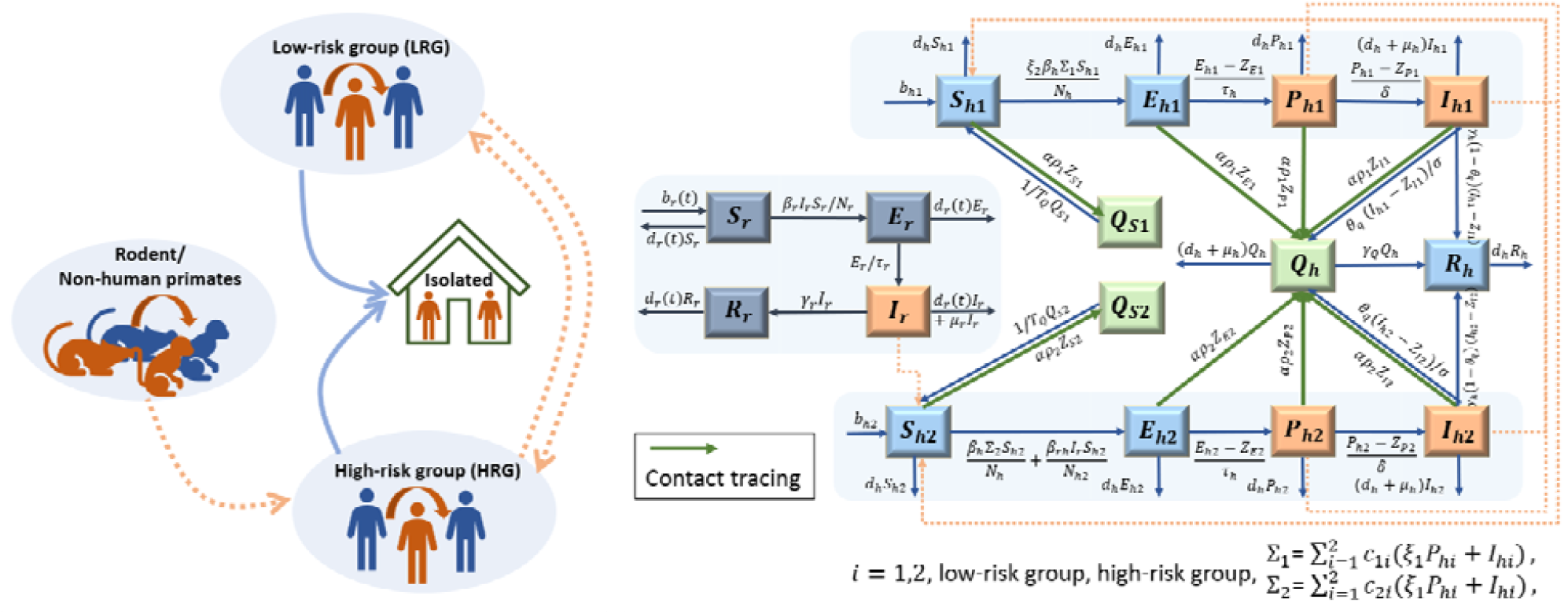
Schematic diagram (left panel) and flow chart (right panel) of the MPX transmission model in both human and rodent populations. The human population is divided into two groups, low-risk group (LRG, i=1) and high-risk group (HRG, i=2).

**Table 1.** Model assumptions.

| Model assumptions |  |
| --- | --- |
| Population classification | ⑦ The human population is divided into two groups, low-risk group (LRG, $i=1$) and high-risk group (HRG, $i=2$). |
| Demographic | ⑦ The birth and death rate of LRG human population and rodent populations are considered. |
| Monkeypox transmission | ⑦ Infectious individuals in both the prodromal phase (with lower infectiousness $\xi_1$, assumed to be 75% of that during the rash stage) and acute phase can transmit the virus.<br>⑦ The traced and isolated individuals are not involved in the transmission.<br>⑦ The transmission risk of LRG individuals is lower than that of HRG individuals, with a scaling factor $\xi_2$ (assumed to be 0.05).<br>⑦ LRG individuals are assumed to have no contact with the rodent population and can only be infected by infectious LRG and HRG individuals.<br>⑦ The HRG individuals can be infected by the infectious LRG and HRG individuals and infectious rodents (By definition, the high-risk group includes individuals who come into contact with rodents because of their work). While the HRG and LRG would equally be exposed to rodents, for simplicity infections in rodents are transmitted to the HRG.<br>⑦ The contacts among LRG individuals mainly come from the LRG individuals, with the proportion $k_1$ (assumed to be 0.6) of the contact rate baseline in the human population.<br>⑦ The contact rate in the HRG is higher than that in the LRG, by a factor $k_2$ of the contact rate baseline (assumed to be 1.3).<br>⑦ The contact between HRG individuals and LRG individuals is low, accounting for $(1 - k_1)$ of the contact rate baseline. |
|  | ☐ The human to rodent transmission is ignored and rodents can only be infected by the infectious rodent.<br>☐ The vertical transmission is ignored. |
| Control measures (if with confirmed case) | ☐ Part of infected individuals with rash symptoms will go to hospital seeking medical help and then be confirmed and isolated.<br>☐ Part of close contact of confirmed symptomatic cases will be traced and quarantined.<br>☐ The traced susceptible people will back to susceptible group after the required quarantined period.<br>☐ Part of the traced individuals will comply with the isolation strategy.<br>☐ The confirmation and isolation of symptomatic case and the tracing and isolation of close contacts requires some time. |

**Table 2.**
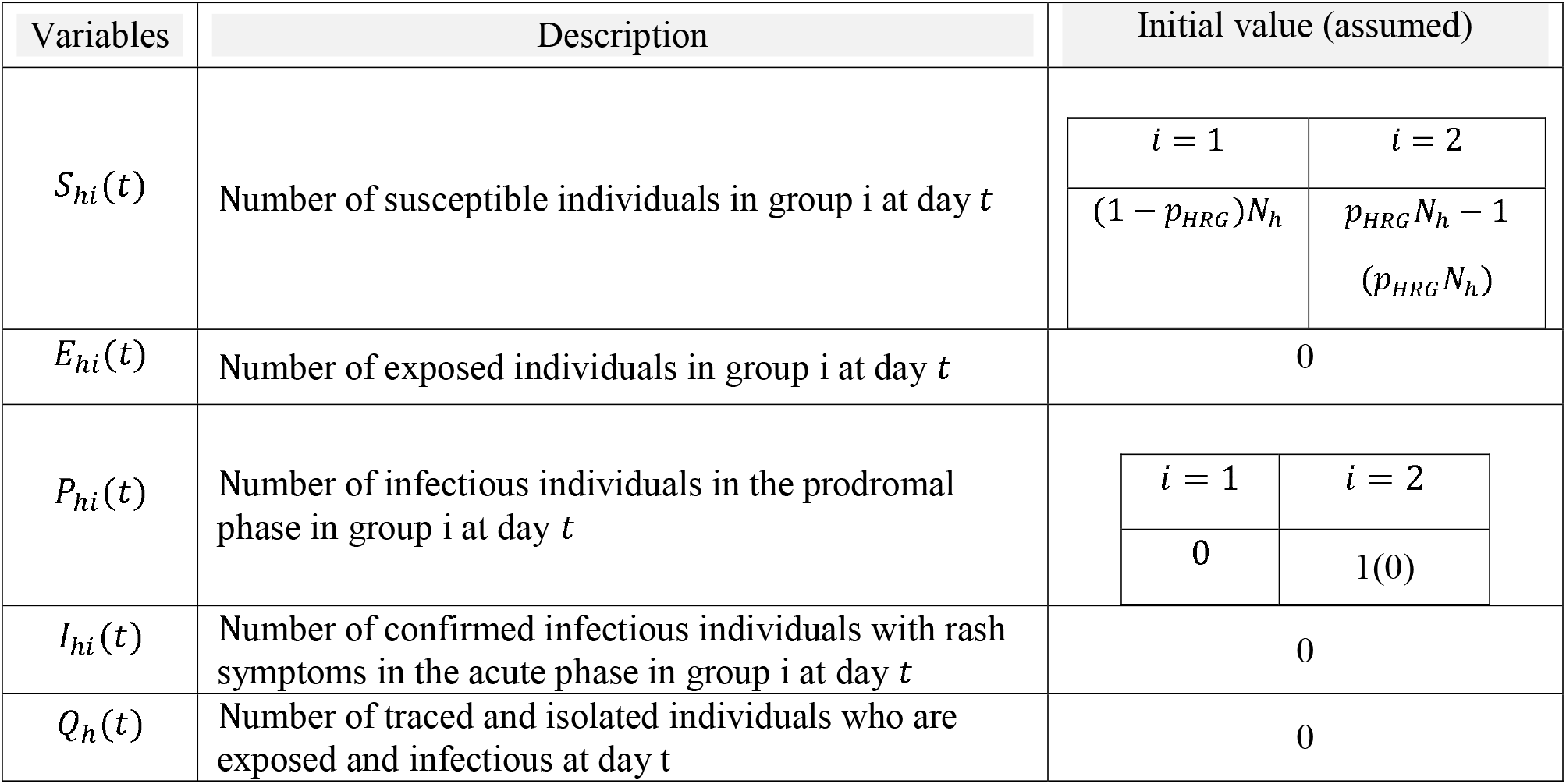
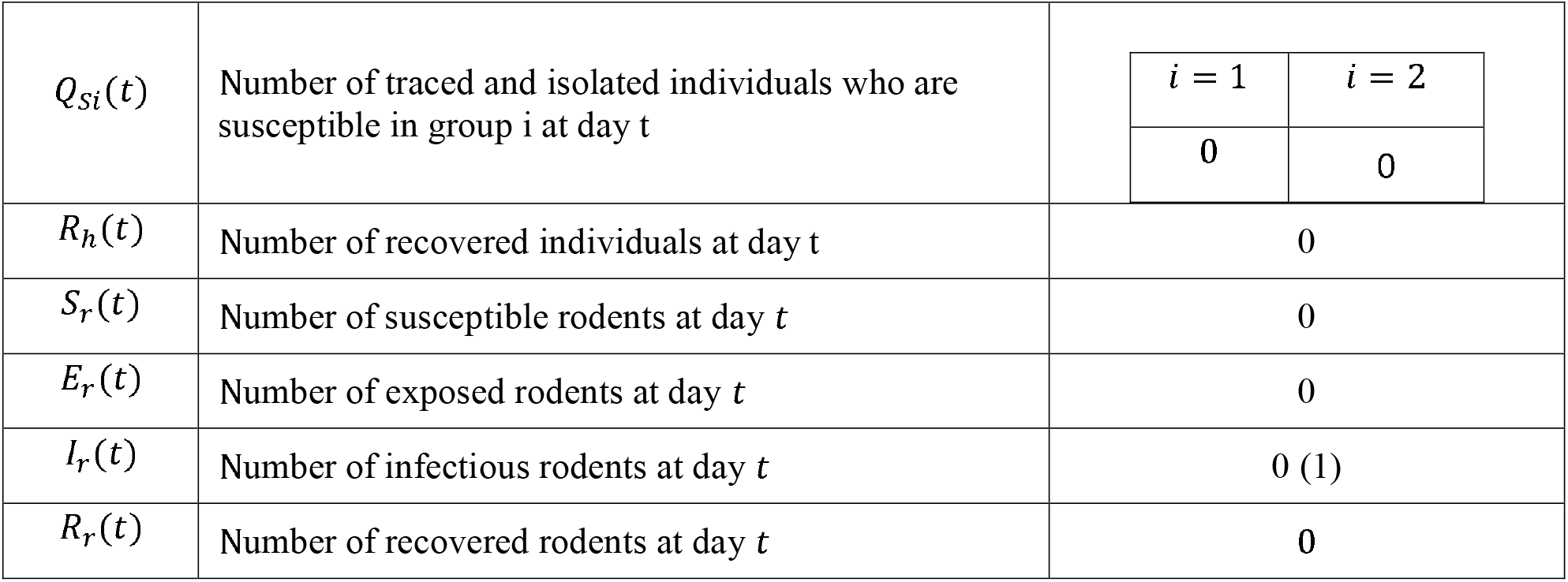
Variables used in the modelling of monkeypox transmission and their assumed initial values.

**Table 3.**
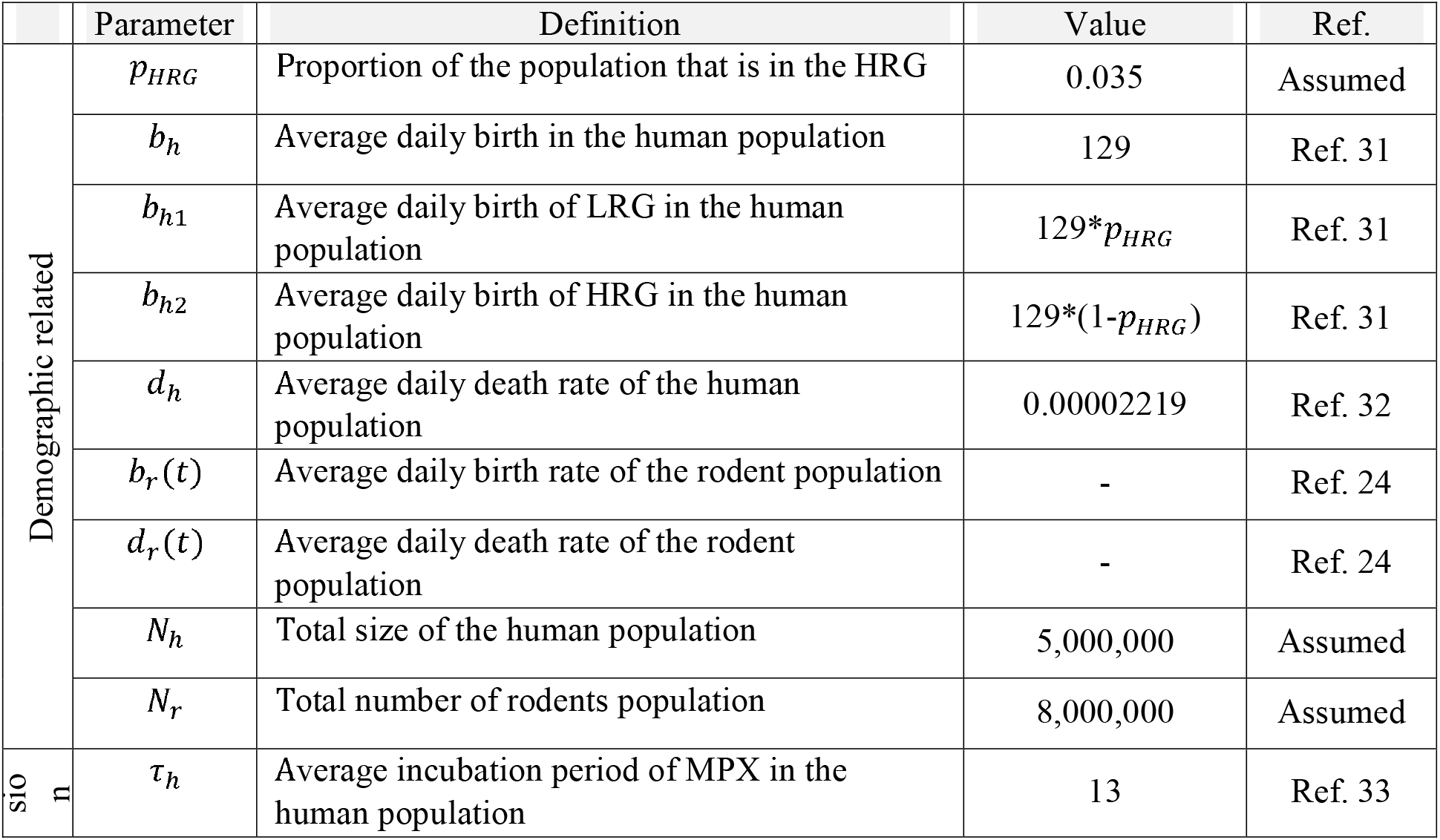
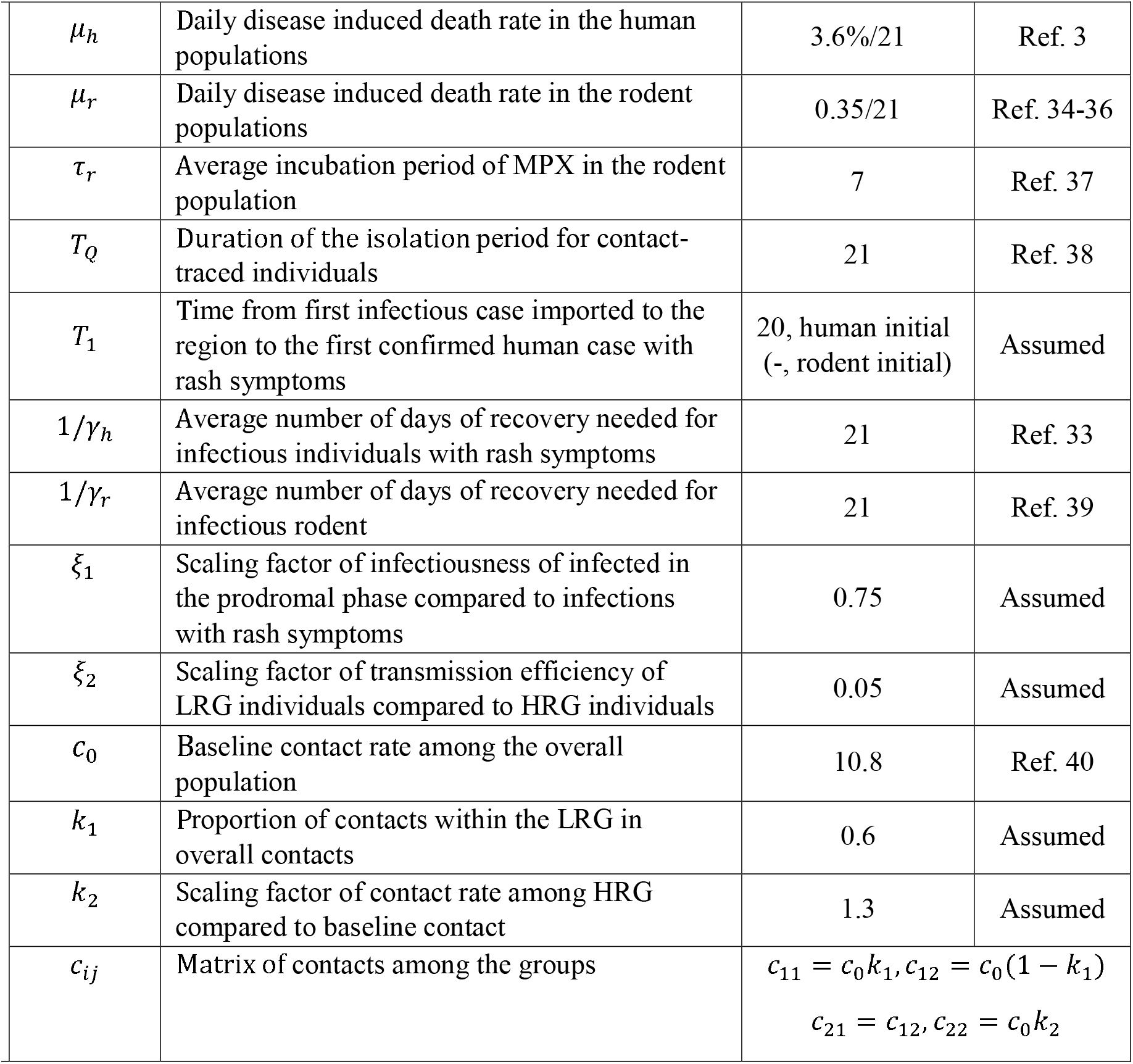
Parameters with the fixed values used in the modelling of monkeypox transmission.

|  | Parameter | Definition | Value | Ref. |
| --- | --- | --- | --- | --- |
| Demographic related | $p_{HRG}$ | Proportion of the population that is in the HRG | 0.035 | Assumed |
| | $b_h$ | Average daily birth in the human population | 129 | Ref. 31 |
| | $b_{h1}$ | Average daily birth of LRG in the human population | $129 * p_{HRG}$ | Ref. 31 |
| | $b_{h2}$ | Average daily birth of HRG in the human population | $129 * (1 - p_{HRG})$ | Ref. 31 |
| | $d_h$ | Average daily death rate of the human population | 0.00002219 | Ref. 32 |
| | $b_r(t)$ | Average daily birth rate of the rodent population | - | Ref. 24 |
| | $d_r(t)$ | Average daily death rate of the rodent population | - | Ref. 24 |
| | $N_h$ | Total size of the human population | 5,000,000 | Assumed |
| | $N_r$ | Total number of rodents population | 8,000,000 | Assumed |
| sio<br>n | $\tau_h$ | Average incubation period of MPX in the human population | 13 | Ref. 33 |

|  |  |  |  |
| --- | --- | --- | --- |
| $\mu_h$ | Daily disease induced death rate in the human populations | 3.6%/21 | Ref. 3 |
| $\mu_r$ | Daily disease induced death rate in the rodent populations | 0.35/21 | Ref. 34-36 |
| $\tau_r$ | Average incubation period of MPX in the rodent population | 7 | Ref. 37 |
| $T_Q$ | Duration of the isolation period for contact-traced individuals | 21 | Ref. 38 |
| $T_1$ | Time from first infectious case imported to the region to the first confirmed human case with rash symptoms | 20, human initial (-, rodent initial) | Assumed |
| $1/\gamma_h$ | Average number of days of recovery needed for infectious individuals with rash symptoms | 21 | Ref. 33 |
| $1/\gamma_r$ | Average number of days of recovery needed for infectious rodent | 21 | Ref. 39 |
| $\xi_1$ | Scaling factor of infectiousness of infected in the prodromal phase compared to infections with rash symptoms | 0.75 | Assumed |
| $\xi_2$ | Scaling factor of transmission efficiency of LRG individuals compared to HRG individuals | 0.05 | Assumed |
| $c_0$ | Baseline contact rate among the overall population | 10.8 | Ref. 40 |
| $k_1$ | Proportion of contacts within the LRG in overall contacts | 0.6 | Assumed |
| $k_2$ | Scaling factor of contact rate among HRG compared to baseline contact | 1.3 | Assumed |
| $c_{ij}$ | Matrix of contacts among the groups | $c_{11} = c_0 k_1, c_{12} = c_0 (1 - k_1)$<br>$c_{21} = c_{12}, c_{22} = c_0 k_2$ | |

**Table 4.** Prior distributions of some key parameters used in the modelling of monkeypox transmission.

| Parameter | Definition | Interval | Source of values |
| --- | --- | --- | --- |
| $\delta$ | Average number of days from prodromal phase to acute phase | 1-5 | Ref. 33 |
| $1/\rho_1$ | Average number of days of the close contact in the LRG needed to be traced | 1-5 | Ref. 2&41 |
| $1/\rho_2$ | Average number of days of the close contacts in the HRG needed to be traced | 1-5 | Ref. 2&41 |
| $\sigma$ | Average number of days for infectious individuals to go from showing rash symptoms to seeking medical help, be confirmed and isolated | 1-13 | Ref. 42 |
| $1/\gamma_Q$ | Average number of days of recovery needed for isolated individuals | 21-36 | Assumed |
| $p_{c1}$ | Percentage of contact tracing in the LRG | 0.1-0.4 | Assumed |
| $p_{c2}$ | Percentage of contact tracing in the HRG | 0.1-0.6 | Assumed |
| $\theta_q$ | Proportion of infectious individuals with rash symptoms who seek medical help, be confirmed and isolated | 0.5-0.8 | Assumed |
| $\alpha$ | Proportion of individuals who comply with the isolation strategy | 0.3-0.5 | Assumed |
| $\beta_h$ | Probability of transmission per contact among HRG | 0.091, 0.114 | Assumed |
| $\beta_r$ | Product of contact rate among rodents and probability of transmission per contact among the rodent population | 0.001, 0.00125 | Assumed |
| $\beta_{rh}$ | Product of contact rate between rodents and humans and probability of transmission per contact from the rodent to the human population | 0.1, 0.125 | Assumed |

### Transmission

Transmission in the human population was modelled using a transmission risk parameter and a contact matrix that describes contacts between and within-population subgroups. The reproduction number of MPX transmission within the HRG was assumed to be 1.2-1.5 for baseline. The reproduction numbers within the HRG and the within the animal reservoir were assumed to be the same. The probability of transmission per contact among HRG was assumed to be 9.1%-11.4%, and 0.46%-0.57% among LRG, respectively. This transmission probability was calculated from the basic reproduction number *R*_0_ derived from our simplified model without public health control measures (see Appendix B for details) by fixing other parameters (Table 3). We also investigated the scenarios with increased reproduction numbers ranging from 1.8-2.25. Moreover, we calculated the control reproduction number *R*_c_ for the original and simplified model with contact tracing and isolation control measures, using the next-generation matrix method (Appendix B)^25^.

### Seasonality of the host rodent population

The seasonality of the host population is thought to be an essential factor affecting MPX transmission^26^. In reservoir host’s breeding season with high activities, the transmission risk of MPX may increase significantly. Here, following the studies of Ogden et al (2007)^24^ we assumed that the rodents have a higher birth rate in the season of spring and summer from April to August, and a higher death rate in the winter period from December to March. Hence, the time-dependent birth rate and death rate of the rodent population were modelled as

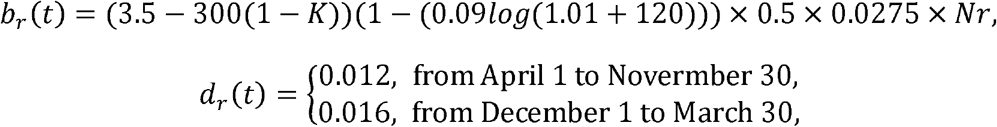

where *Nr* is the total number of rodents, and

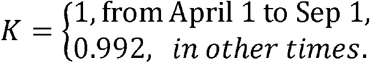

Here, we considered a normal season for rodent population reproduction, and the situation of more favourable conditions for rodent reproduction due to climate change is not considered. The seasonality of the population dynamics of rodents was also presented in Appendix **Figure 1**.

### Scenario analysis

We performed numerical simulations of our model with the setting of a hypothetical metropolitan city with initial human and rodent population sizes of 5,000,000 and 8,000,000, respectively. The starting time of the simulation was set as May 1, 2022; the model is then run for 1000 days. Tables 2, 3, and 4 give the initial values, parameters with fixed values, and prior distribution of some parameters used for simulations, respectively. Table 5 lists and describes all the scenarios projecting the daily new infections in the LRG, HRG, and rodent populations (per 100k) presented. In scenarios A, B, C, the mean, and confidence interval of infection cases are obtained from 5000 parameter sets sampling from the prior distribution (uniform) of parameters, by the Latin hypercube sampling (LHS) method^27^. In scenarios D and E, we only presented the mean of all the simulations. We conducted analyses using MATLAB (R2020a)^28^.

**Table 5.** List of scenarios’ settings used to project cases of monkeypox.

| Scenarios | Settings |
| --- | --- |
| Scenario A | A human infectious case is imported into the region. The proportion of individuals in the HRG among the total population is 3.5%.<br>☐ No public health control measures are conducted (**Baseline**).<br>☐ No contact tracing, 65% of infections with rash symptoms are isolated after confirmation. |
| Scenario B | A human infectious case is imported into the region. The transmission efficiency among human population increases by 50%. The proportion of individuals in the HRG among the total population is 3.5%.<br>☐ No contact tracing, 65% of infections with rash symptoms are isolated after confirmation.<br>☐ 65% of infections with rash symptoms are isolated after confirmation, and contact tracing concerns 60% of close contacts of infections in the HRG. |
| Scenario C | A rodent infectious case is imported into the region.<br>☐ No public health control measures are conducted.<br>☐ No contact tracing, 65% of infections with rash symptoms are isolated after confirmation.<br>☐ The overall transmission efficiency increases by 50%. 65% of infections with rash symptoms are isolated after confirmation and contact tracing concerns 60% of close contacts of infections in the HRG. |
| Scenario D | A human infectious case is imported into the region. The transmission efficiency among human population increases by 50%. 65% infections with rash symptoms are isolated after confirmation. The proportion of individuals in the HRG among the total population is 3.5%.<br>☐ Contact tracing 30%/ 50%/ 70% of close contact of infections in the HRG.<br>☐ Contact tracing 30%/ 50%/ 70% of close contact of infections in the LRG. |
| Scenario F | A human infectious case is imported into the region. No contact tracing. 65% of infections with rash symptoms are isolated after confirmation.<br>☐ The proportion of individuals in the HRG among the total population is 2% / 4% / 6%. |

### Sensitivity analysis and key factors

We conducted a sensitivity analysis of key parameters to address the uncertainty of parameters, by employing the LHS and partial rank correlation coefficient (PRCC) method^27^. We generated 2000 samples of these investigated parameters, including the probability of transmission per contact among HRG, the isolation proportion of symptomatic cases, the proportion of contact tracing in the LRG and HRG, and the number of rodents in the initial state. The ranges of parameters used in the LHS are reported in Appendix Table A1. We then explored the relationship between the parameters and the cumulative cases by calculating the values of PRCC; parameters with a PRCC magnitude above□0.5 are considered to be significant in the model outcomes^29^. In addition, we conducted the sensitivity analysis regarding the control reproduction number *R*_*c*_ for the original model without simplification using the parameters associated with the control measures and compared the effect of different control measures by normalising the correlation coefficient.

## Results

### Isolation of infectious cases to control monkeypox spread

An outbreak of MPX transmission is possible if public health measures are not in place, even when only human-to-human transmission is involved (**Figure 2A**). The average peak of daily new infections of MPX in HRG may reach 100 for the metropolitan area with HRG individuals accounting for 2% if no public health control measures are implemented. However, this risk can be greatly reduced if an isolation strategy is implemented. If 65% of the infectious cases are isolated, MPX transmission can be controlled or maintained at a very low level of endemicity (**Figure 2B**). For a metropolitan area with 5,000,000 residents, the average number of daily new infections in the HRG is below 1 under the assumption of *R*_0_ between 1.2 and 1.5, if the isolation strategy is respected efficiently.

**Figure 2.**
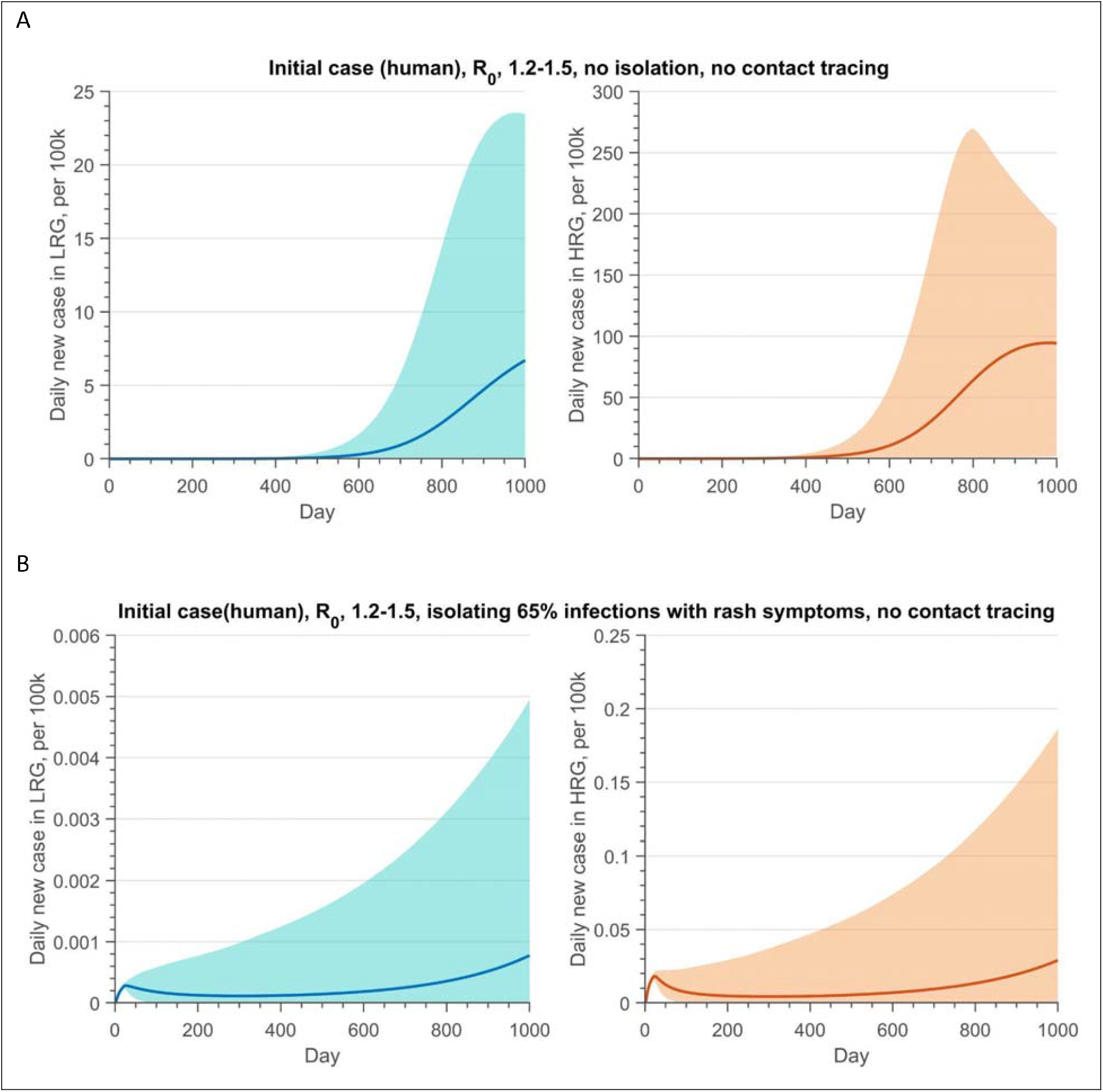
Projections of daily new infections (per 100k) in the LRG and HRG if the initial case is a human infection, under the different public health control measures. Panel A, no isolation, and no contact tracing; panel B, no contact tracing but isolation of 65% of infections with rash symptoms.

### Impact and concerns about possible increased transmission efficiency

There is evidence of increased transmission efficiency among gbMSM^30^. Hence, we investigate the scenario that the transmission risk increases by 50%, making the within HRG *R*_0_ ranged between 1.8 and 2.25. Under this hypothesis, the average number of daily new MPX cases reaches 200 per 100k in HRG and 100 per 100k in LRG despite 65% of infections with rash being isolated (**Figure 3A**). However, the cases drop significantly in both high-risk and low-risk groups of people if 60% of the close contact of infection can be traced along with the 65% rash-motivated isolation. And the peak will arrive much later. This indicates that tracing close contacts of infection with rash in HRG will be an efficient way to reduce the infection size among the human population facing a possible increased transmission efficiency of the virus (**Figure 3B, 5**). Besides, we conducted the sensitivity analysis of the transmission risk among the human population to justify the parameter uncertainty (**Figure 7**).

**Figure 3.**
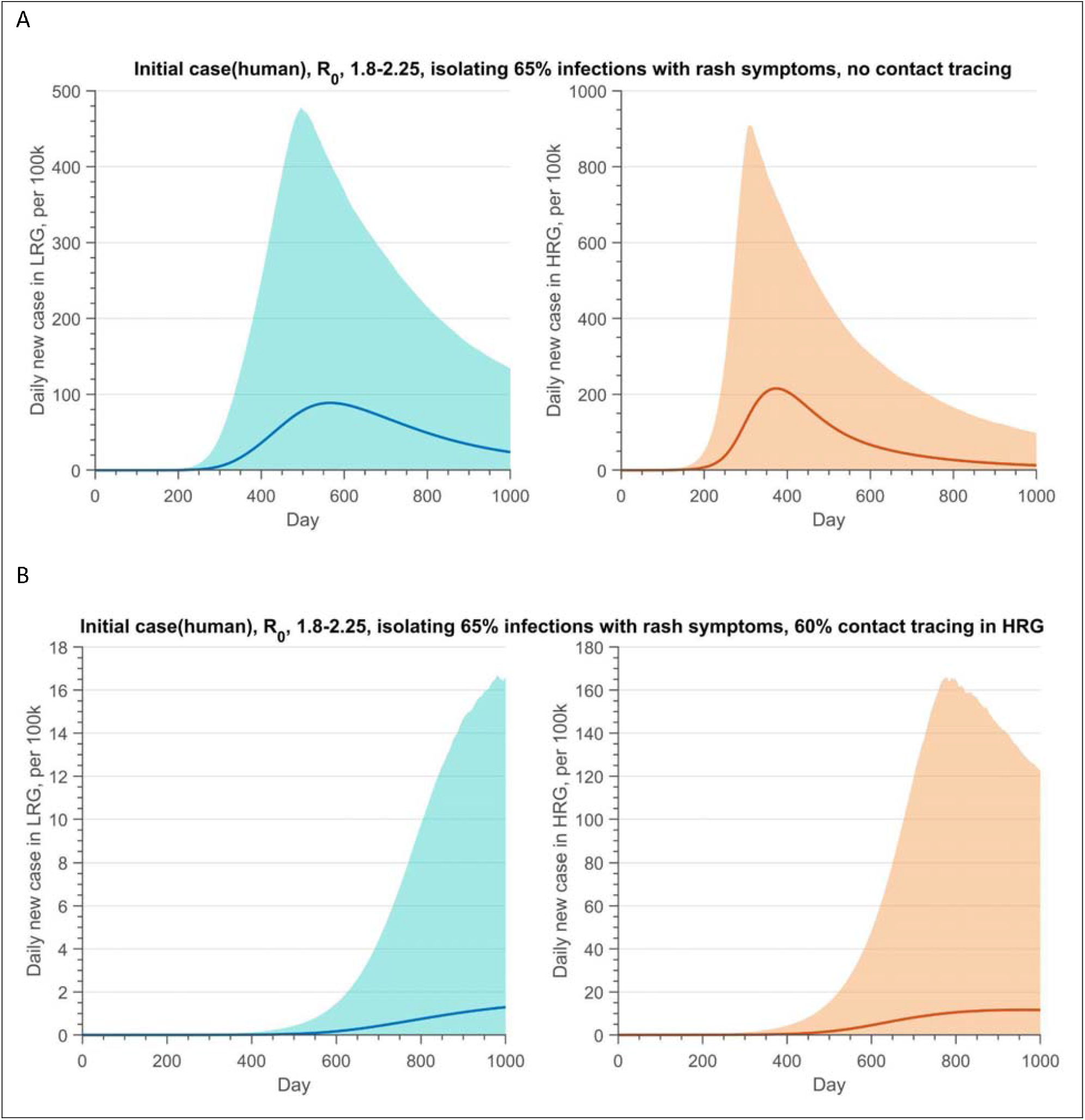
Projections of daily new infections (per 100k) in the LRG and HRG if the initial case is a human infection and the transmission efficiency increases by 50%, under the different public health control measures. Panel A, no contact tracing and isolation of 65% of infections with rash symptoms; panel B, isolating 65% of infections with rash symptoms and tracing 60% of close contacts of the infections with rash symptoms in the HRG.

### The role of animals in monkeypox transmission

Humans are at greater risk of outbreaks if animal hosts contribute to the transmission (**Figure 4**). We also observe that rash-motivated isolation may not be sufficient to protect the susceptible HRG individuals due to the reinforcement of animal-to-human transmission, although the total number of cases decreases (**Figure 4B**). Moreover, the situation can become even more worrisome under the possibility of increased transmission efficiency due to the evolution of the virus and the incidence of host and environment. We may observe much earlier peaks and multiple waves driven by animal transmission (**Figure 4C**). The isolation strategy is beneficial for epidemic mitigation (**Figure 7**), nevertheless, it may not be sufficient if the virus spreads in the animal populations. Other public health measures such as contact tracing, monitoring, and controlling infections in the animal population, may also be needed.

**Figure 4.**
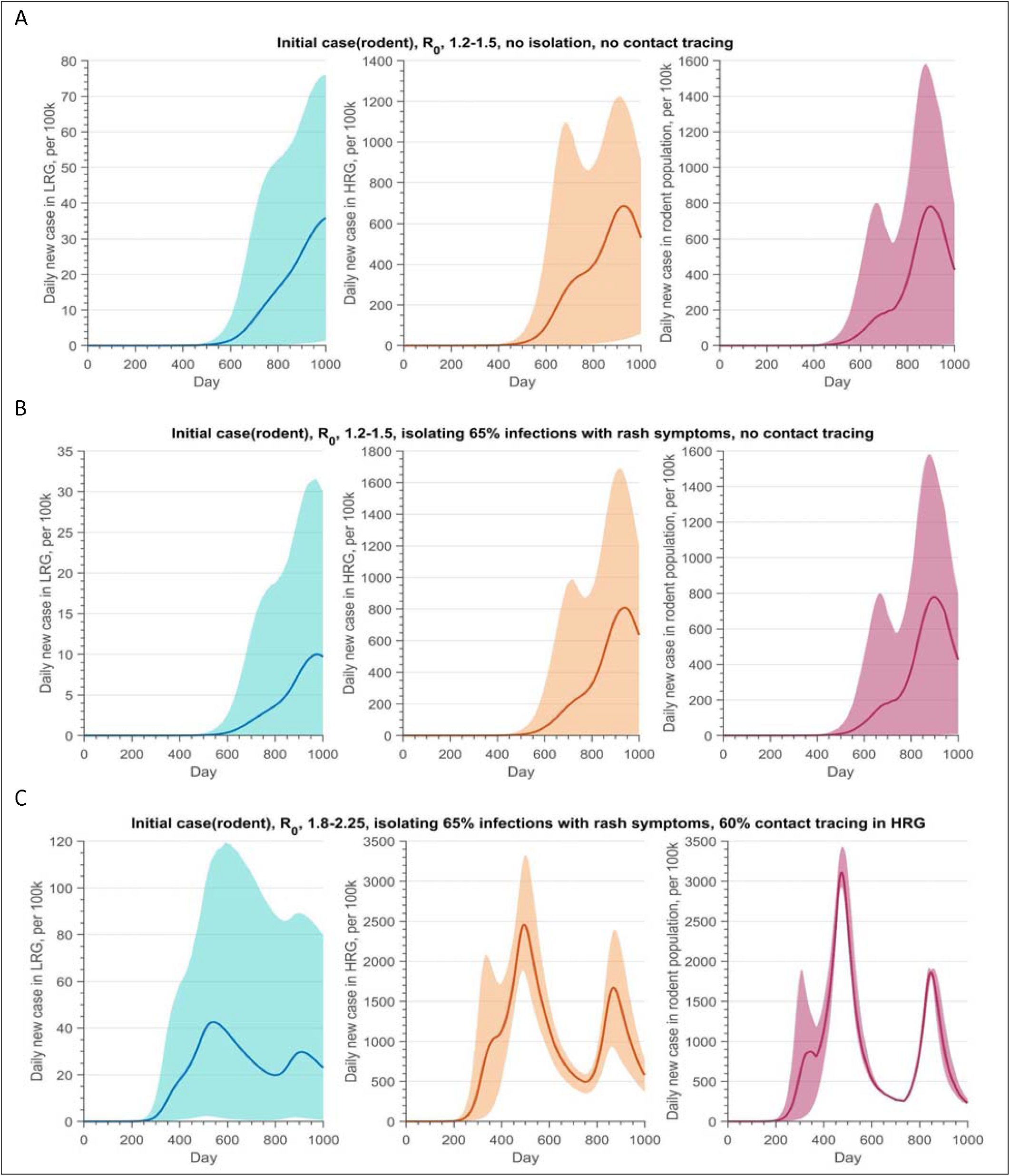
Projections of daily new infections (per 100k) in the LRG, HRG, and rodent population if rodents are involved in the transmission under different scenarios. Panel A, no isolation, and no contact tracing; Panel B, isolating 65% of infections with rash symptoms and no contact tracing; panel C, isolating 65% of infections with rash symptoms and tracing 60% of close contacts of the infections with rash symptoms in the HRG and increased 50% of overall transmission efficiency.

### Contact tracing in HRG helps to control monkeypox spread

Figure 5 presents the comparison of mitigation effects on MPX transmission among contact tracing in HRG and LRG, under the assumption of *R*_0_ between 1.8 and 2.25 and 65% rash-motivated isolation. Contact tracing in HRG shows a better effect on the containment of transmission as expected, compared to tracing in LRG. In addition, it shows the possibility of keeping the MPX prevalence at a low level by employing control measures in combination with rash-motivated isolation and contacting tracing in HRG, even facing the 50% increased transmission efficiency.

**Figure 5.**
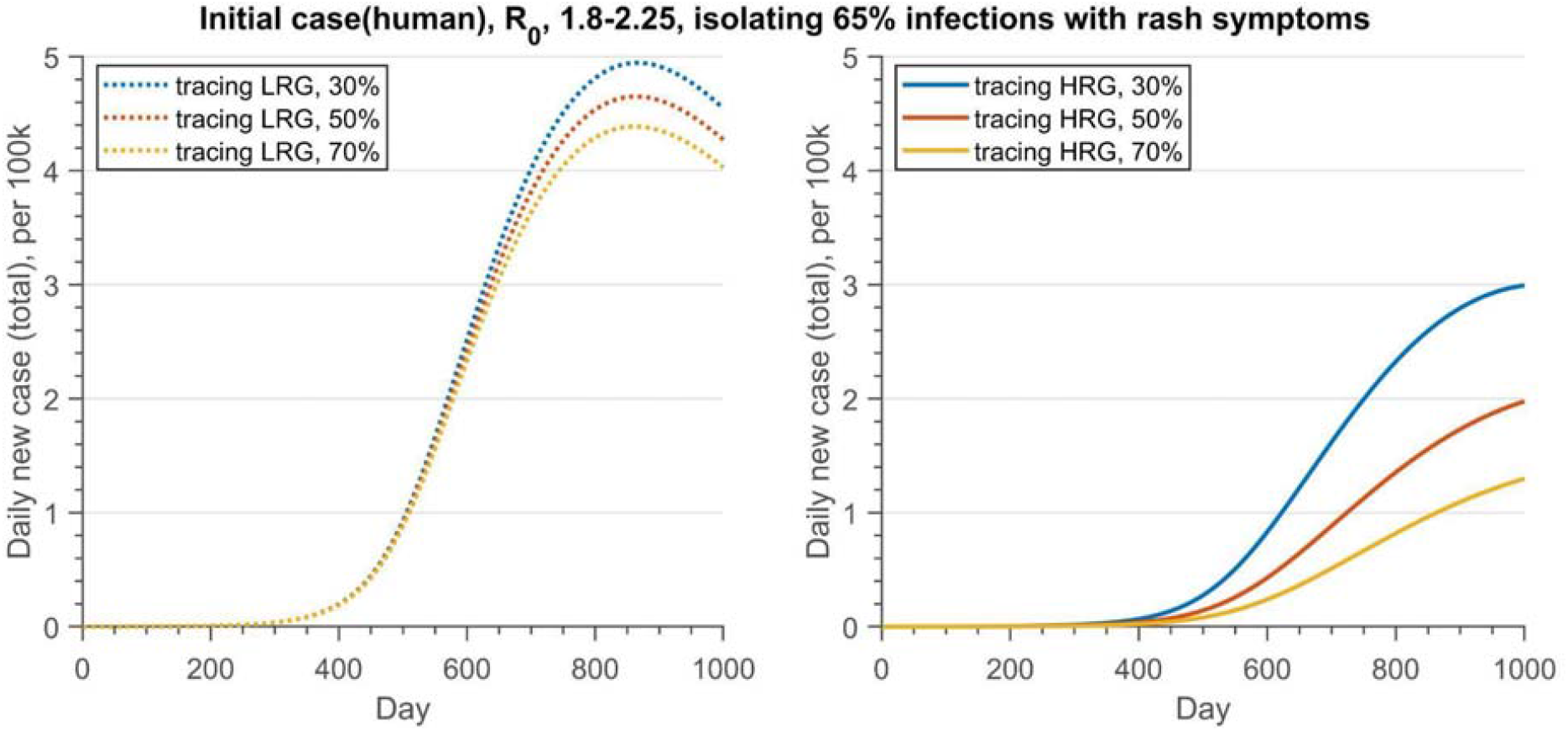
Projections of daily new infections (per 100k) in the LRG and HRG if the transmission efficiency increases by 50% and 65% of infections with rash symptoms are isolated, under the different contact tracing proportions in the HRG (solid line) and LRG (dash line), 30% (light blue), 50% (orange) and 70% (yellow). Note that when the contact tracing in HRG is included, there is no contact tracing in LRG, and vice versa.

### Regions with larger number of gbMSM communities need more attention

Moreover, the smaller the initial proportion of individuals in the high-risk group, the fewer infections in the human population (**Figure 6**), which is also confirmed by the sensitivity analysis (**Figure 7**). This result suggests that in a community with the proportion of HRG being 2 or 4%, it takes a longer time before seeing cases start increasing, whereas a 6% increase of the size of the group leads to a much earlier peak. This result indicates the importance of educating the public about MPX transmission, since a city with a higher population of gbMSM has a much higher possibility of a MPX outbreak.

**Figure 6.**
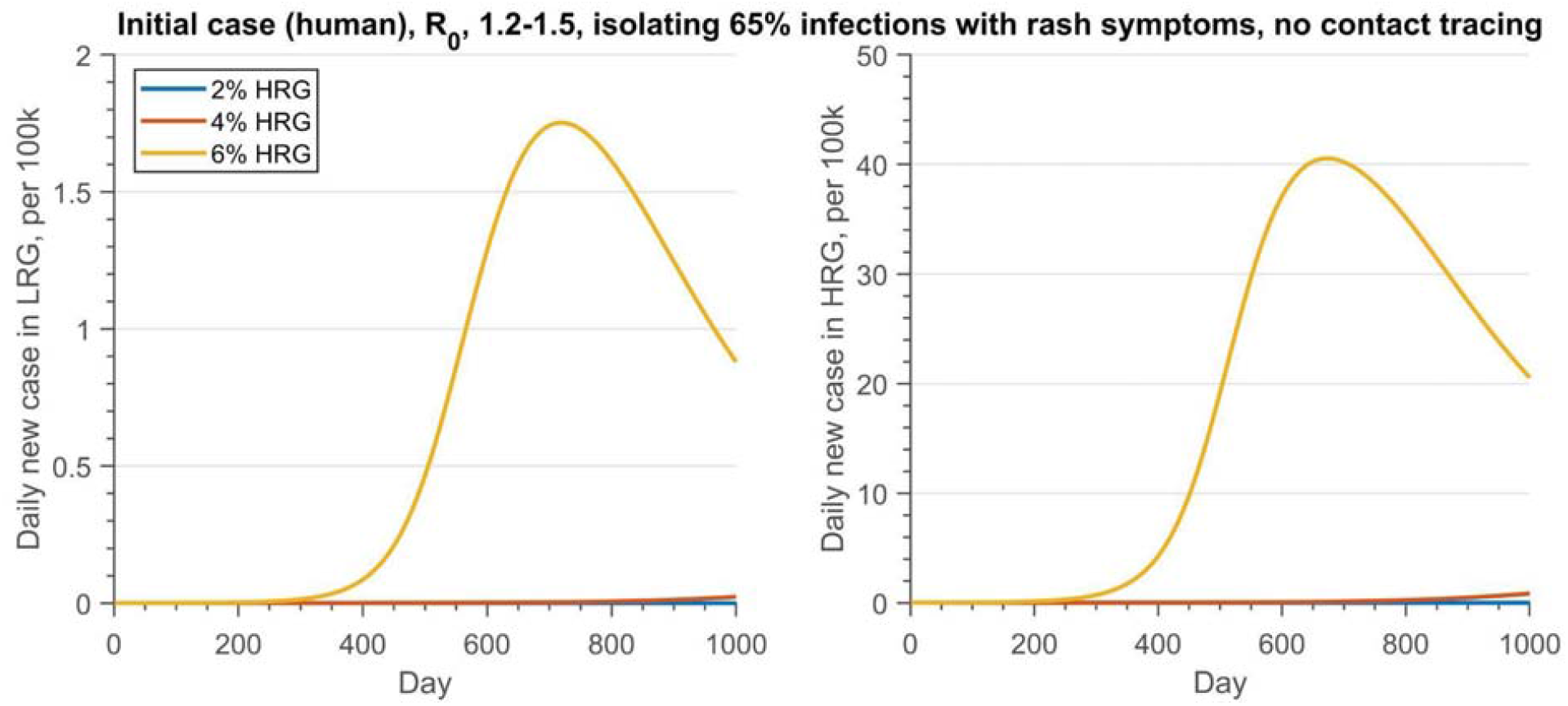
Projections of daily new infections (per 100k) in the LRG and HRG if isolating 65% of infections with rash symptoms, under the different initial proportions of HRG individuals, 2% (light blue), 4% (orange) and 6% (yellow).

**Figure 7.**
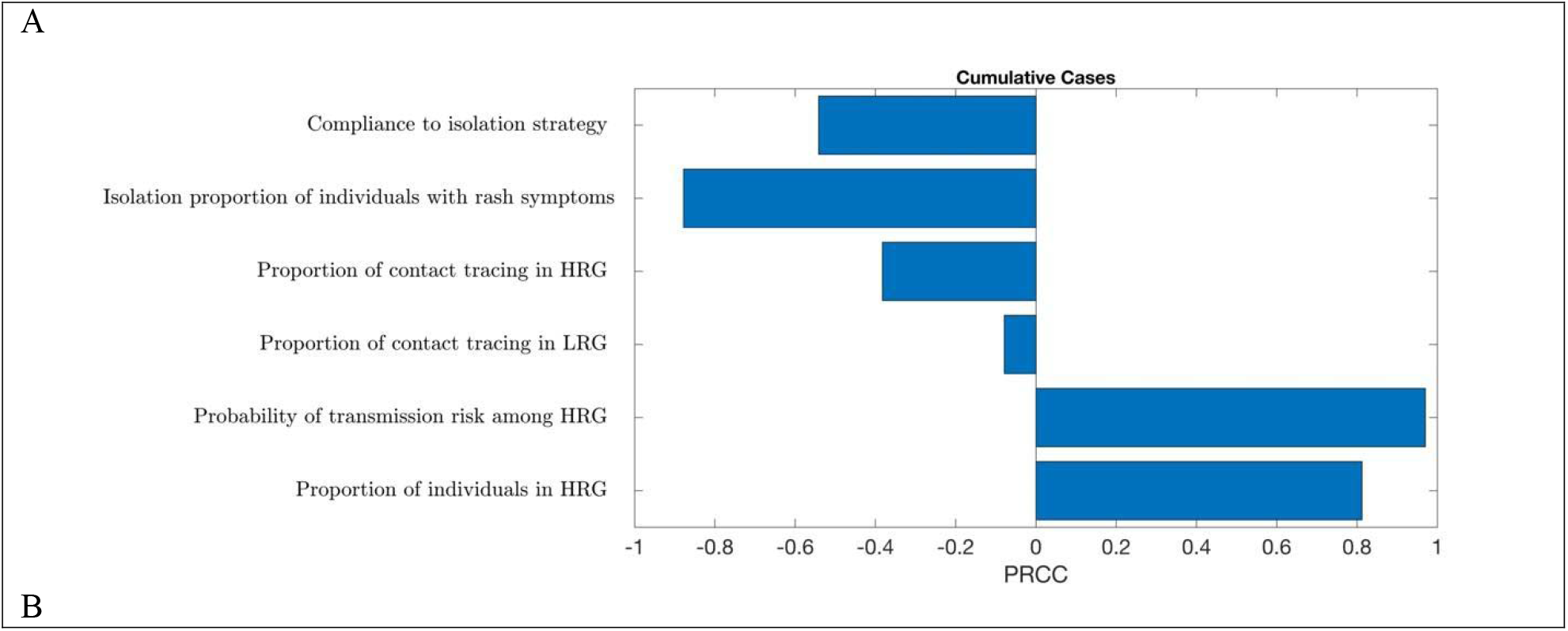
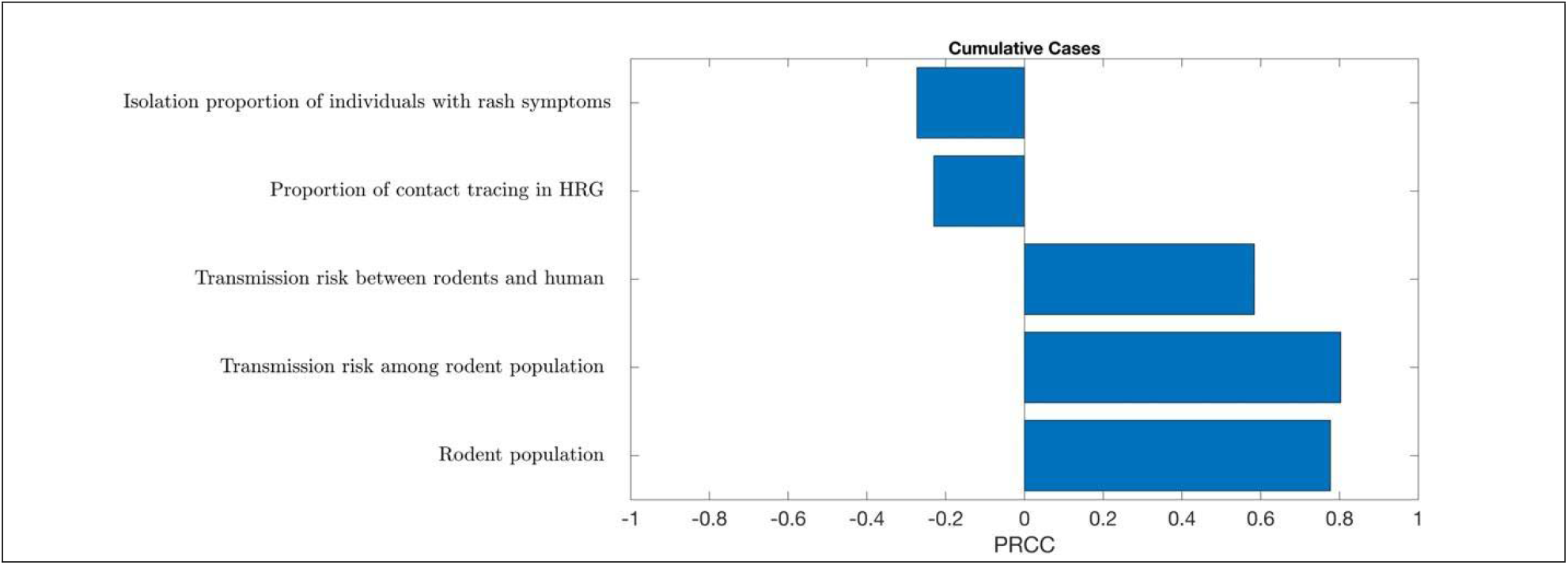
The sensitivity analysis on cumulative cases if the initial case is a human infection (A) or rodent infection (B).

### Sensitivity analysis and key factors

We observe that HRG individuals have a significant effect on the cumulative cases. Our results also show that rash-motivated case isolation presents a negative correlation with cumulative cases **(Figure 7A)**. Meanwhile, compliance with the isolation strategy and implementation of contact tracing in HRG are negatively correlated to the cumulative cases, indicating the importance of those control measurements in mitigating the MPX outbreak **(Figure 7A)**. On the other hand, if the animal populations are involved in the MPX transmission, the cumulative number of human cases is distinctly influenced by the size of the rodent population, the transmission risk among the rodent population, and therefore risk between rodents and humans **(Figure 7B)**. The sensitivity analysis suggests that control of MPX requires not only rash-motivated isolation, but also efficient contact tracing, and population adherence to the strategy. Furthermore, the monitoring and controlling of the infection among the animal populations is also crucial to protect the broader human population.

**Figure 8** shows that the most significant measure is the rash-motivated isolation strategy. Meanwhile, the average number of days for infectious individuals from showing rash symptoms to seeking medical help then to be confirmed and isolated also play a key role in the prevention and control of the disease. Moreover, the contact tracing in HRG is more important than that in LRG, which is also illustrated by the sensitivity analysis of cumulative cases.

**Figure 8.**
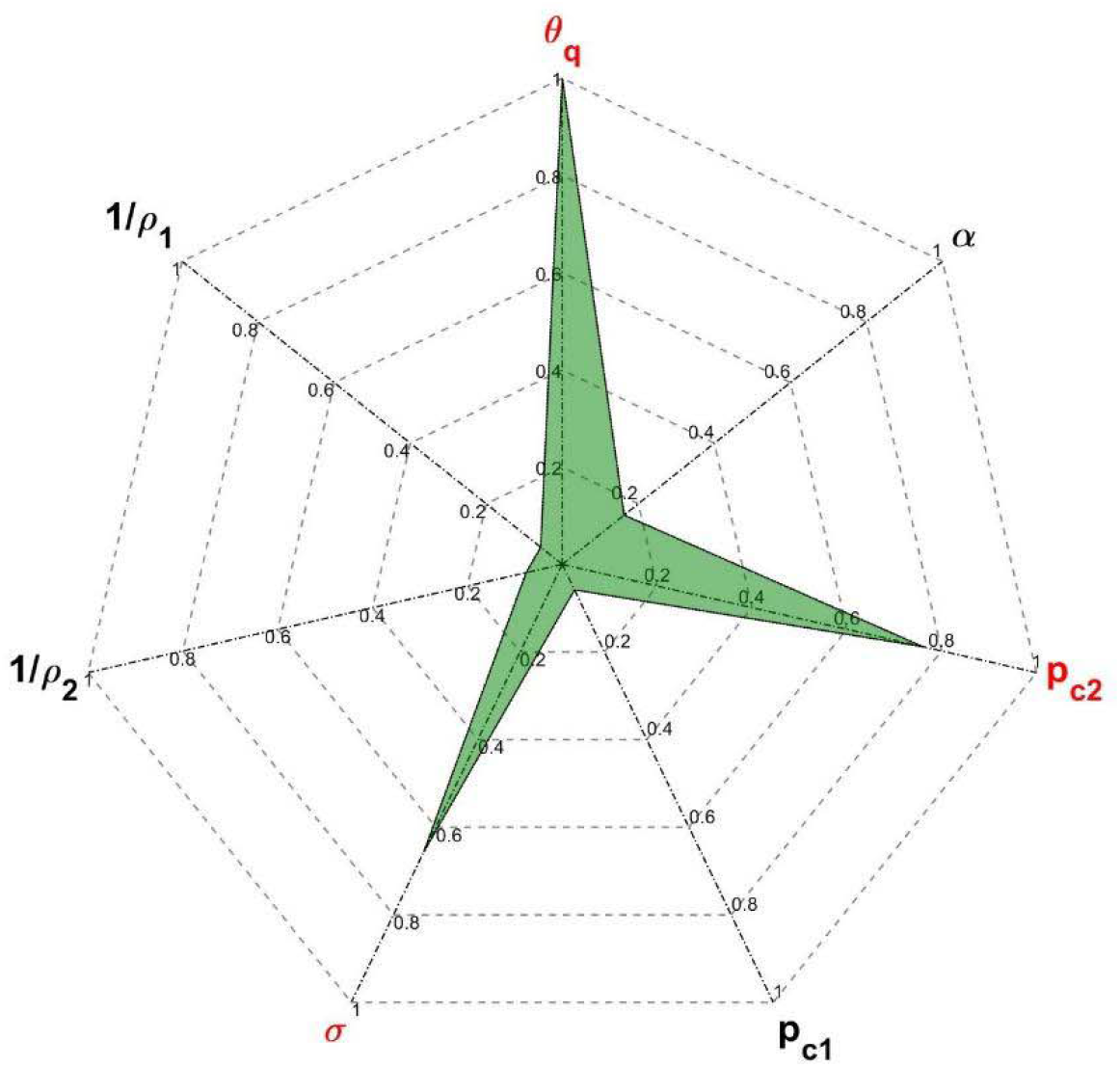
The radar diagram of sensitivity analysis regarding control reproduction number *R*_*c*_ without simplification with the control measures. *θ*_*q*_ is the proportion of infectious individuals with rash symptoms who seek medical help and then be confirmed and isolated; *σ* represents the average number of days for infectious individuals from showing rash symptoms to seek medical help (then be confirmed and isolated); *p*_*c*1_ (*p*_*c*2_) is the percentage of contact tracing in the LRG (HRG); 1/*ρ*_1_ (1/*ρ*_2_) is the average number of days of the close contact in the LRG (HRG) needed to be traced and isolated; *α* is the proportion of individuals who comply with the isolation strategy. The notations in red color indicate that the P-value of the parameter is significant.

## Discussions

Cases of MPX have been rising worldwide in the past months, raising widespread concern. Since Monkeypox is a zoonosis, we have used a One Health approach to model MPX transmission and assess the risk of outbreak in an hypothetical metropolitan area inhabited by humans of different risk groups, and animal hosts as well. We have also conducted a sensitivity analysis to identify the key parameters needed to establish confidence in our model, notwithstanding the uncertainties in current MPX transmission.

Our simulations suggest that the risk of a MPX outbreak remains high, especially in high-risk groups in the absence of intervention. In particular, the general population may be at risk if transmission efficiency increases among gbMSM or other HRGs. Such an occurrence due to virus evolution could render isolation of symptomatic cases insufficient to stop the spreading, and case detection and contact tracing in HRG would become an essential strategy to prevent bridging from HRG to LRG. Furthermore, regions with a higher proportion of gbMSM may face higher risks and require more stringent public health measures, including isolation and contact tracing. Moreover, the spread of the virus being closely associated with seasonal variations in animal host abundance and activity level, it is vital to monitor the incidence of MPX in animals to serve as an indicator for assessing the risk of an MPX epidemics.

Although contact tracing in the gbMSM community is fraught with a number of ethical considerations concerning privacy, such as sexual behavior, education campaigns and frank communication are important to detect higher risks of infection^10^. In this context, the risk of stigmatising gbMSM cannot be underestimated if the control measures (like vaccines) focus on the gbMSM, and the consequences in delaying the seeking of medical care in these communities. On the other hand, raising awareness of the risk of MPX transmission, and ways to reduce contacts and risks of exposure to infection, is also crucial.

Our modelling studies support the latest assessment of the World Health Organization that the current spreading of MPX does not constitute a global health emergency^5^. However, our take home message is that the public health administration should be on high alert and take a more aggressive public health response when local emerging cases start, just like what the US CDC is doing to activate emergency operation centres for MPX response^43^.

Our modelling scenario simulations utilise parameters from available literature for previous outbreaks in Africa, and could thus be improved with the e accumulation of data in newly emerging areas, allowing more refined estimations of key model parameters to inform public health decision making. In the meantime, we will be able to determine the minimum required proportion of contact tracing to prevent MPX outbreaks. In our model we did not consider the increasing concern of human-to-animal transmission routes since no such cases have been documented, and the risk is estimated to be low. The vaccination strategy may be applied to the high-risk groups, and thus need to be considered in our future work too.

## Data Availability

All data produced in the present work are contained in the manuscript

## Author Contributions

Conceptualization: H.Z., P.Y., Y.T., L.Y., NO; Data curation: Y.T., L.Y., P.Y.; Formal analysis, L.Y., P.Y., Y.T., H.Z.; Methodology: P.Y., Y.T., L.Y., H.Z.; Software: P.Y., Y.T., L.Y.; Validation: P.Y., Y.T., L.Y.; Visualization: P.Y., Y.T., L.Y.; Writing - original draft: P.Y., Y.T., L.Y., E.A., H.Z.; Writing - review & editing: J.A., J.B., J.H., J.W., N.O., H.B., E.A., P.Y., Y.T., L.Y., H.Z.; Funding acquisition: H.Z., J.A., J.B., J.H., J.W., H.B.; Supervision: H.Z.

## Appendix

### Appendix A Compartment model

We constructed a deterministic compartmental model of MPX transmission within and between human and animal populations. The human population was divided into two groups: the low-risk group (LRG, i.e. broader population [subscript 1]) and the high-risk group (HRG, i.e. members of the gbMSM community with multiple sexual partners [subscript 2]). An animal population (with [subscript *r*]) is included in the model, which could comprise any of the mentioned potential reservoir species, as well as new reservoirs if they were to emerge. For simplicity, we refer to the animal population as the rodent population in the simulations. The model follows the SEIR framework, with the extension including Infectious *P* (prodromal phase) – Infectious *I* (rash phase) – Isolated *Q*_*h*_ (infectious) – Isolated *Q*_*s*_ (susceptible) subpopulations (**Figure 1**). A description of model assumptions, variables and parameters used are presented in Tables 1-4.

Transmission occurs as follows: susceptible individuals become infected at rate *β*_*h*_*c*_*ij*_, where *β*_*h*_ is the probability of transmission per effective contact (The transmission risk of LRG individuals is lower than that of HRG individuals, with a scaling factor *ξ*_2_) and *c*_*ij*_ (*i,j* = 1,2)is the average daily contacts among subgroups (indicating the average daily contacts that people in group *i* have with people in group *j*), after encountering infectious individuals in the prodromal phase, with lower transmission risk, *ξ*_1_, and rash phases. As shown in Figure 1, after an incubation period, (*τ*_*h*_), infected individuals become infectious in the prodromal phases and after a period, *δ*, develop a rash and eventually recover at rate *γ*_*h*_, or a proportion *θ*_*q*_ is isolated after being confirmed in *σ* days. Through contact tracing at rate *ρ*_1_, the close contacts of confirmed infections, including susceptible, exposed, and infected individuals, may also be detected and isolated. It is recognized that quarantine of contacts would be practically difficult due to the long (up to 4 weeks) infectious period, but post-exposure vaccination of contacts may provide a kind of “effective quarantine”. We assume their compliance to the isolation policy is *α*, which is the product of the proportion of people who adhere to the isolation protocol and their compliance time. Contact traced susceptible individuals will be isolated for *T*_*Q*_ days, after which they return to the susceptible compartment. Contact traced exposed and infected individuals may recover at the same rate *γ*_*Q*_ or die at a rate *µ*_*h*_, for simplicity. LRG individuals may face a lower infection risk compared to HRG individuals, with a scaling factor *ξ*_2_. By definition, the high-risk group includes individuals who come into contact with rodents because of their work; so HRG individuals may also be infected by contacts with infected rodents at a rate *β*_*rh*_. Infections in the HRG may receive more attention, hence we assume a different contact tracing rate *ρ*_2_ for them. Susceptible rodents are infected by infectious rodents at rate *β*_*r*_, and after the incubation period (*τ*_*r*_), exposed rodents will become infectious and then recover at rate *γ*_*r*_, or die at rate *µ*_*r*_. Also, natural birth and death rates of the human and rodent populations are considered. The daily new birth of humans is *b*_*h*_ and the natural death rate is *d*_*h*_ The seasonality of the population dynamics of rodents is included in the model, with birth rate *b*_*r*_(*t*) and natural death rate *d*_*r*_(*t*).

#### The contact tracing process

We denote 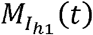 and 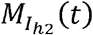 as the daily new confirmed monkeypox cases in the LRG and HRG, respectively. They are determined as follows, where *σ* is the average number of days for infectious individuals from showing rash symptoms to seeking medical treatment, be confirmed and isolated:

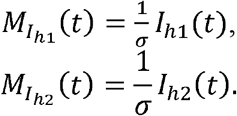

The number of traced close contacts of newly confirmed cases in LRG (*Z*_1_) and HRG (*Z*_2_) at time *t* are

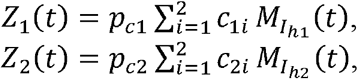

where *p*_*c*1_ and *p*_*c*2_ are the percentage of contact tracing in the LRG and HRG respectively. We assume that 0 ≤ *p*_*ci*_ ≤ 1, *i* = 1,2.

The traced close contacts are composed of the susceptible, exposed, and infectious (prodromal and rash phases) individuals. The close contacts of confirmed cases are proportionally distributed in the LRG and HRG based on the contact matrix. Applying a similar idea as used in Yuan et al. (2022)^1^, we obtain the following number of traced individuals in the LRG that are exposed (denoted as *Z*_*E*1_(*t*)):

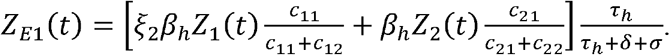

Similarly, we can obtain the number of traced infections in prodromal phase and rash phase in the LRG, as

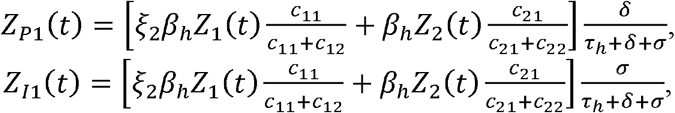

respectively.

The number of traced individuals in the LRG that are susceptible is

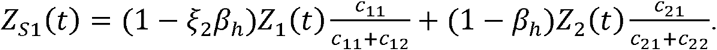

Hence, the traced susceptible, exposed, and infectious (prodromal phase and rash phases) individuals in the HRG are

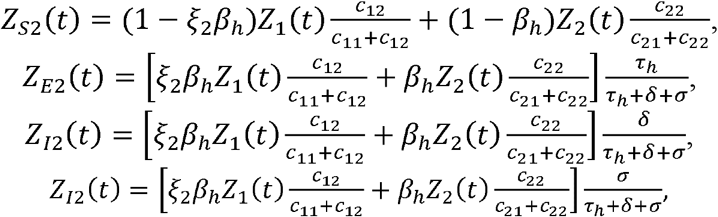

respectively.

#### Model equations

Following the flowchart in Figure 1, the transmission of monkeypox in a metropolitan area is described by the following system of differential equations

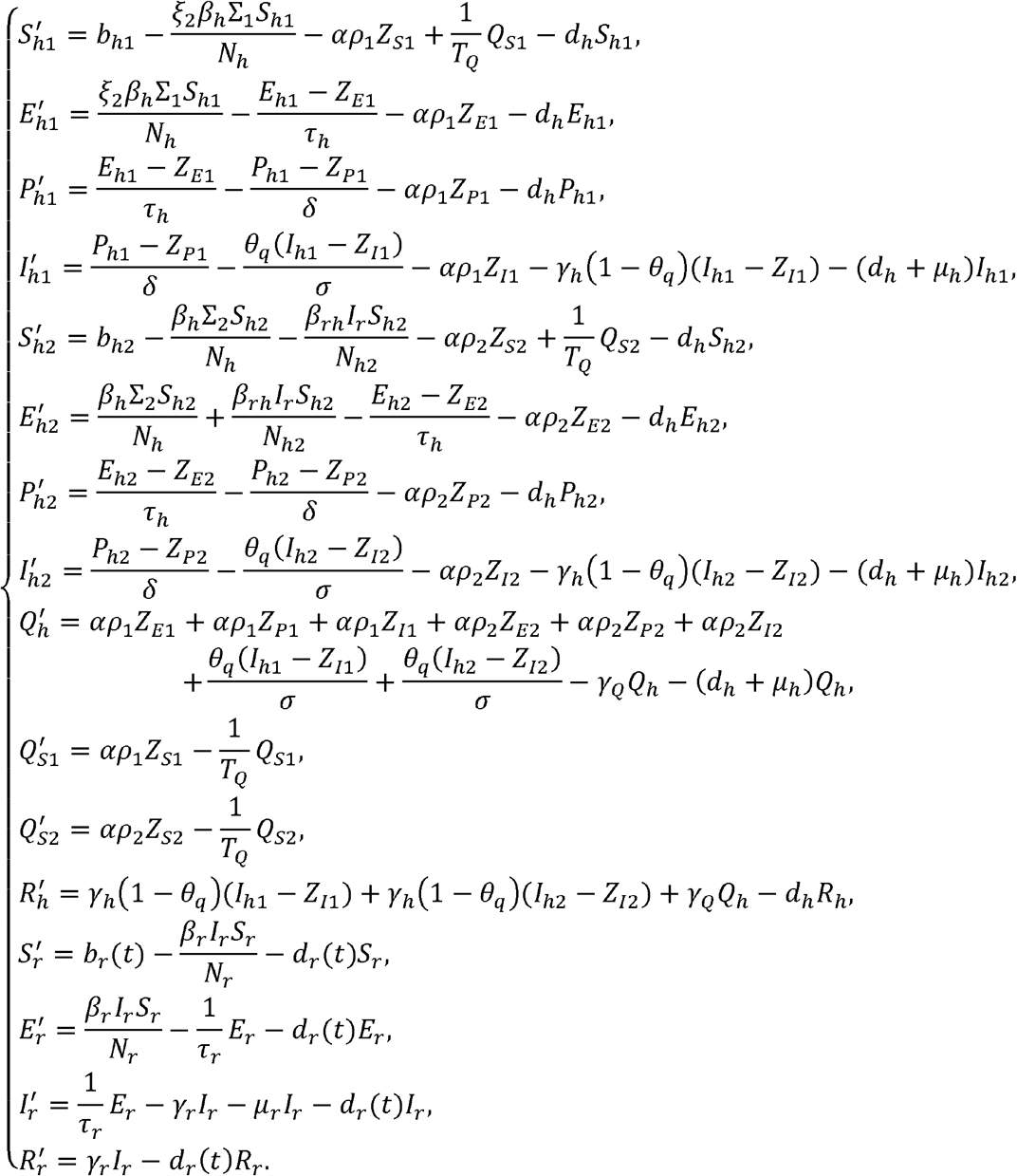

where 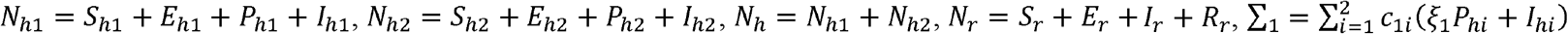 and 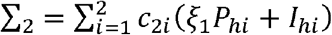.

If there is no isolation and contact tracing, then we have the simplified model

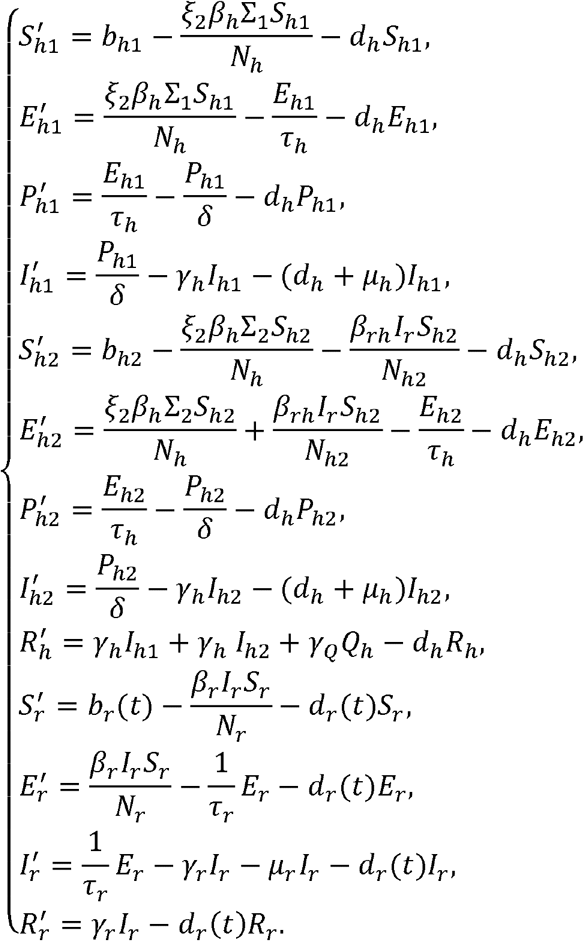

where 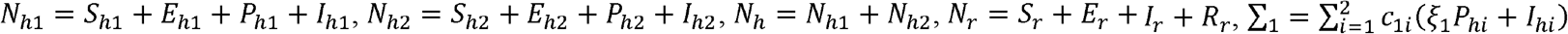 and 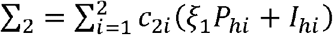.

### Appendix B Basic and control reproduction number

Let the birth rate term “*b*_*r*_(*t*)” and death rate term “*d*_*r*_(*t*)” be constants, denoted “*b*_*r*_” and “*d*_*r*_” respectively. For the simplified model without public health control measures, we calculated the basic reproduction number following the next generation matrix method, yielding

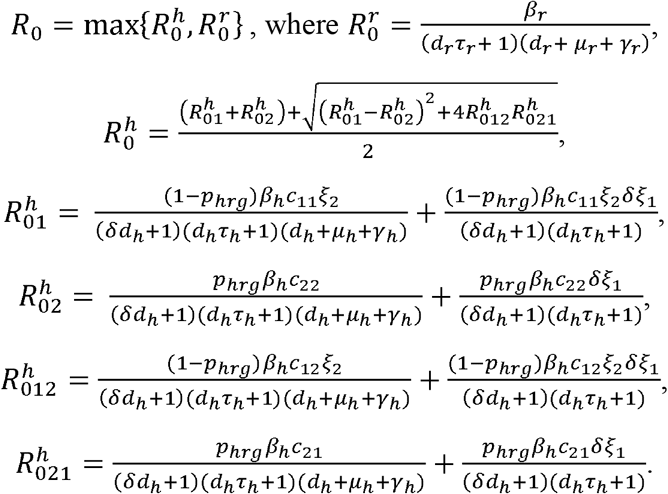

For the model with public health control measures with rash-motivated isolation and contact tracing, we also obtain the control reproduction number *R*_*c*_ using the next generation matrix method.

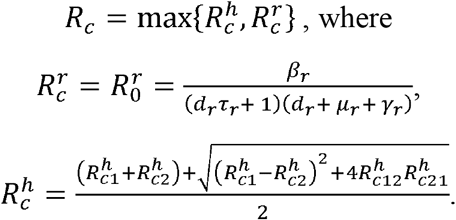

Definitions of 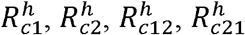 are in the Appendix.

#### The coefficients in control reproduction number of the original model

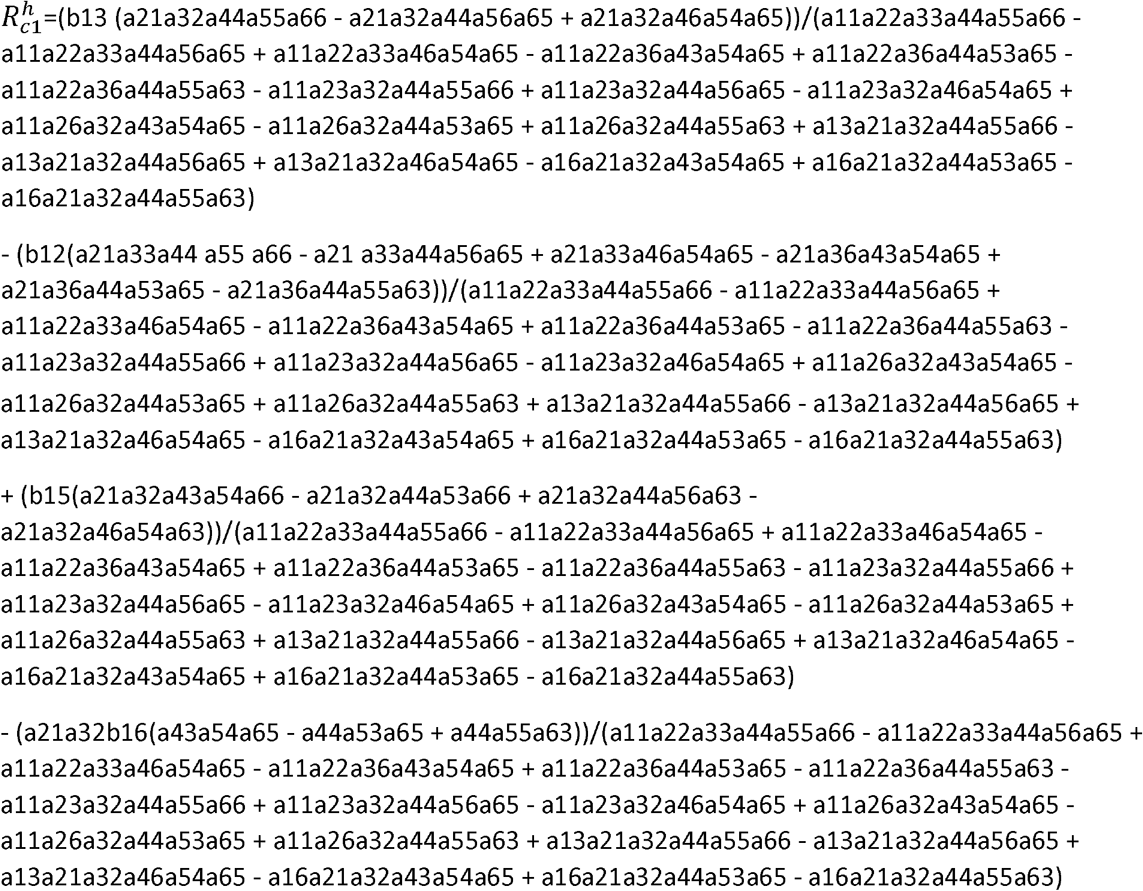

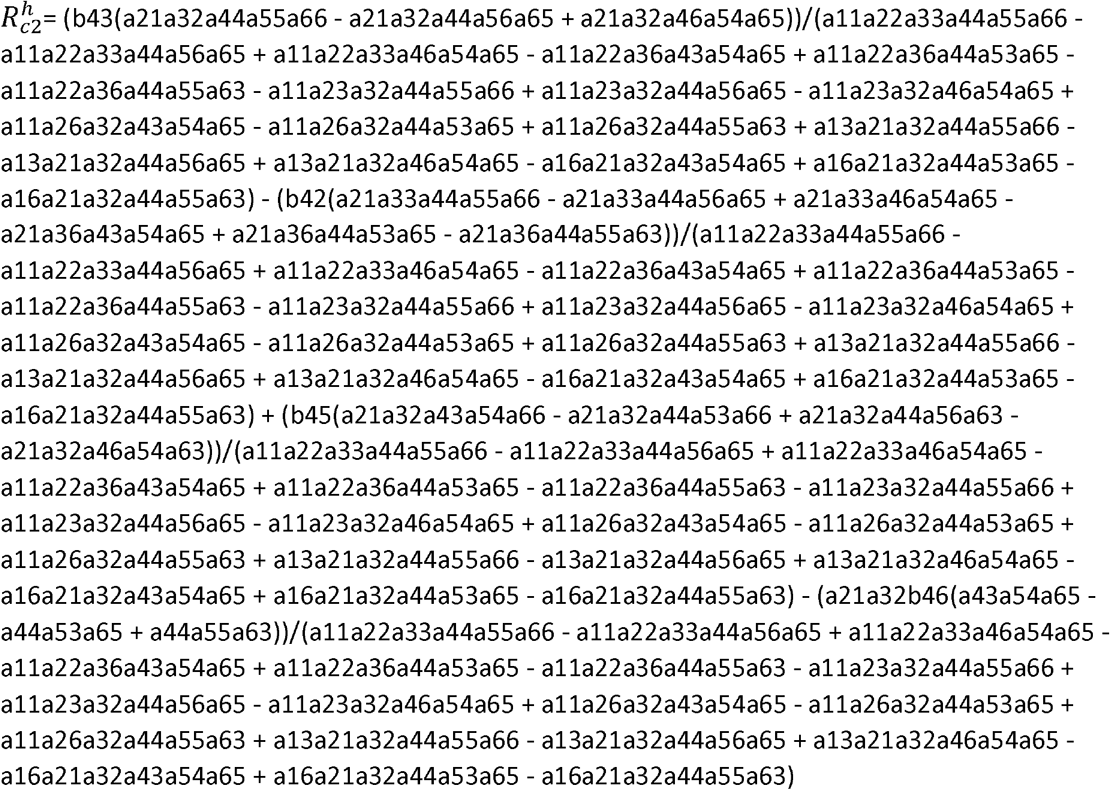

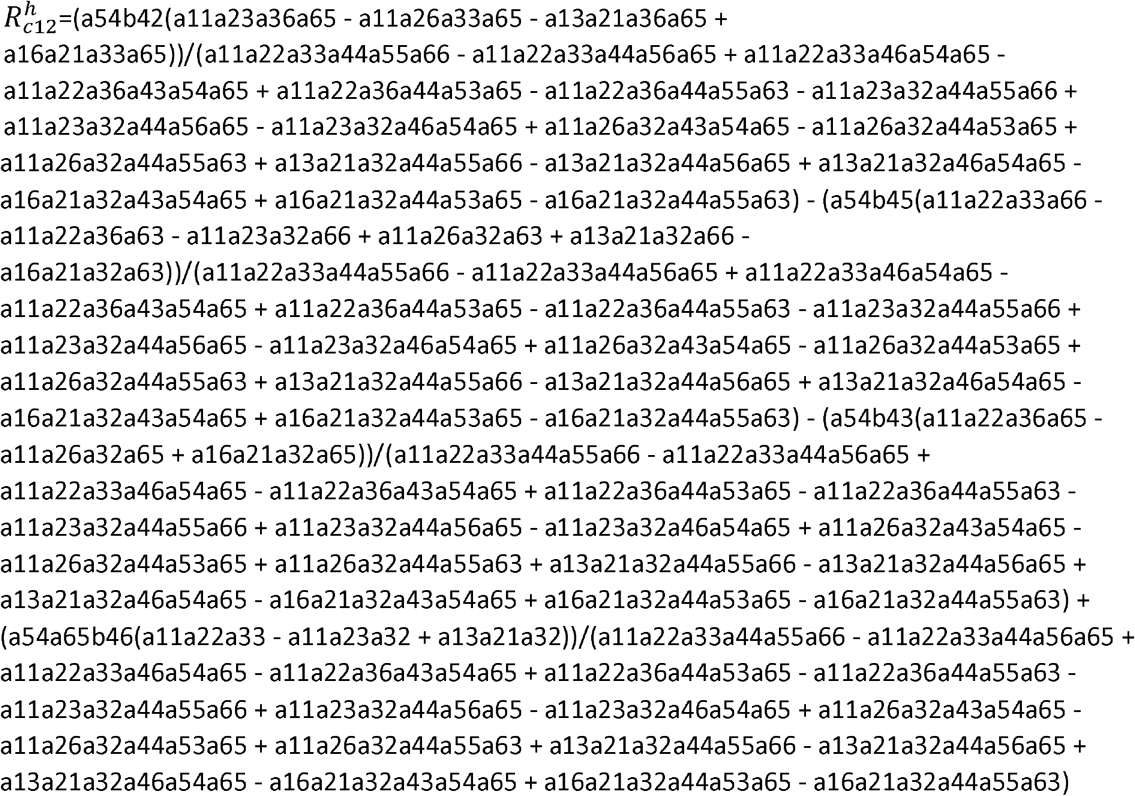

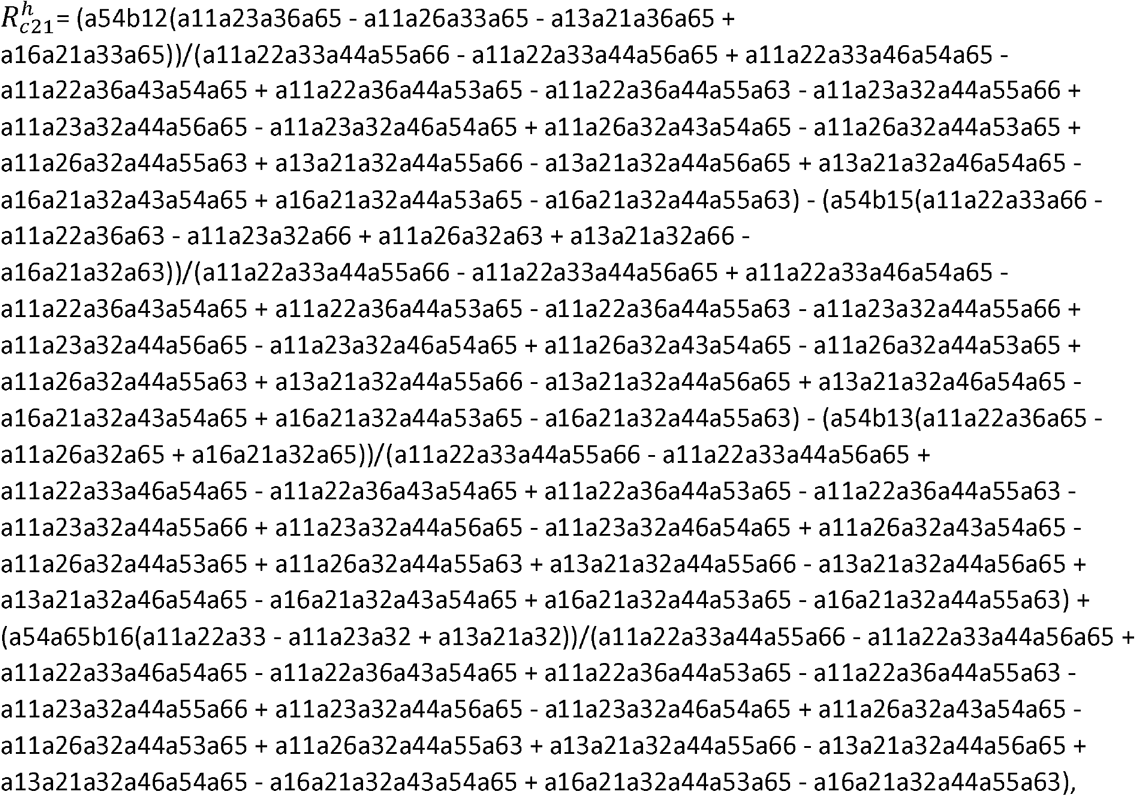

where

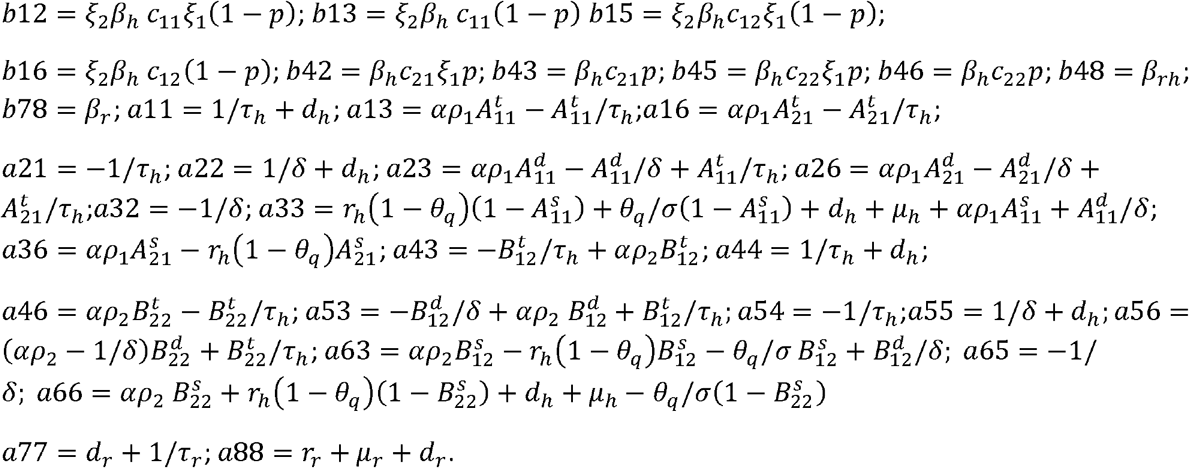

Here we denote

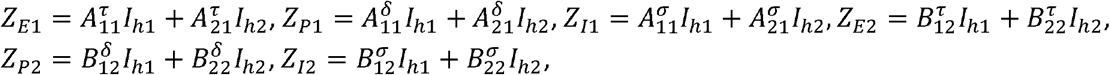

where 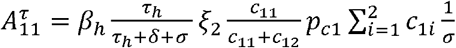,

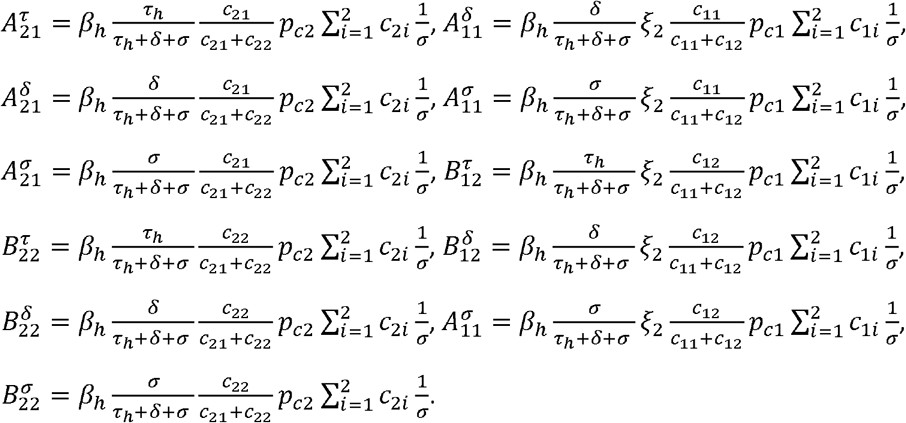

To better understand how the public health control measure affect the control reproduction number, we simplify the contact tracing terms “*αρ*_1_*Z*_*E*1_, *αρ*_1_*Z*_*p*1_, *αρ*_1_*Z*_*I*1_, *αρ*_2_*Z*_*E*2_, *αρ*_2_*Z*_*p*2_, *αρ*_2_*Z*_*I*2_” as “*η*_*E*1_*E*_*h*1_, *η*_*p*1_*P*_*h*1_, *η*_*I*1_*I*_*h*1_, *η*_*E*2_*E*_*h*2_, *η*_*P*2_*P*_*h2*_, *η*_*E*1_*E*_*h*1_, *η*_*I*2_*I*_*h2*_”, respectively, where *η*_*i*_, *i* = *E*l, *P*l, *I*1, *E*2, *P*2,*I*2, denotes by isolation rate for those infectious people being traced. It is the combined factors including the proportion of compliance to the isolation strategy, the days needed for tracing and the percentage of contact tracing. Then the control reproduction number becomes

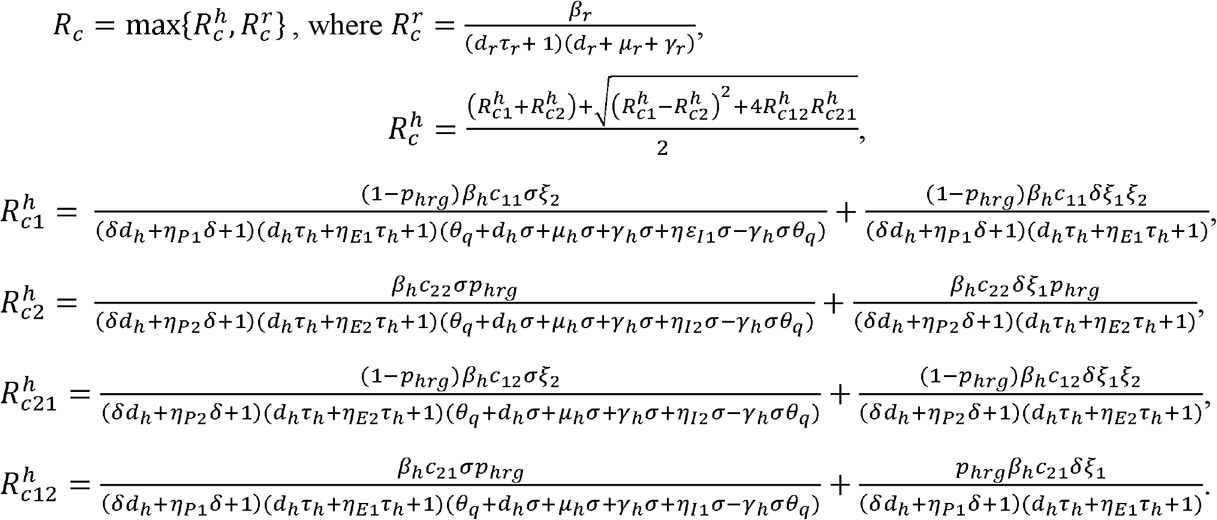

### Appendix C Seasonality dynamics of the rodent population

**Figure A1:**
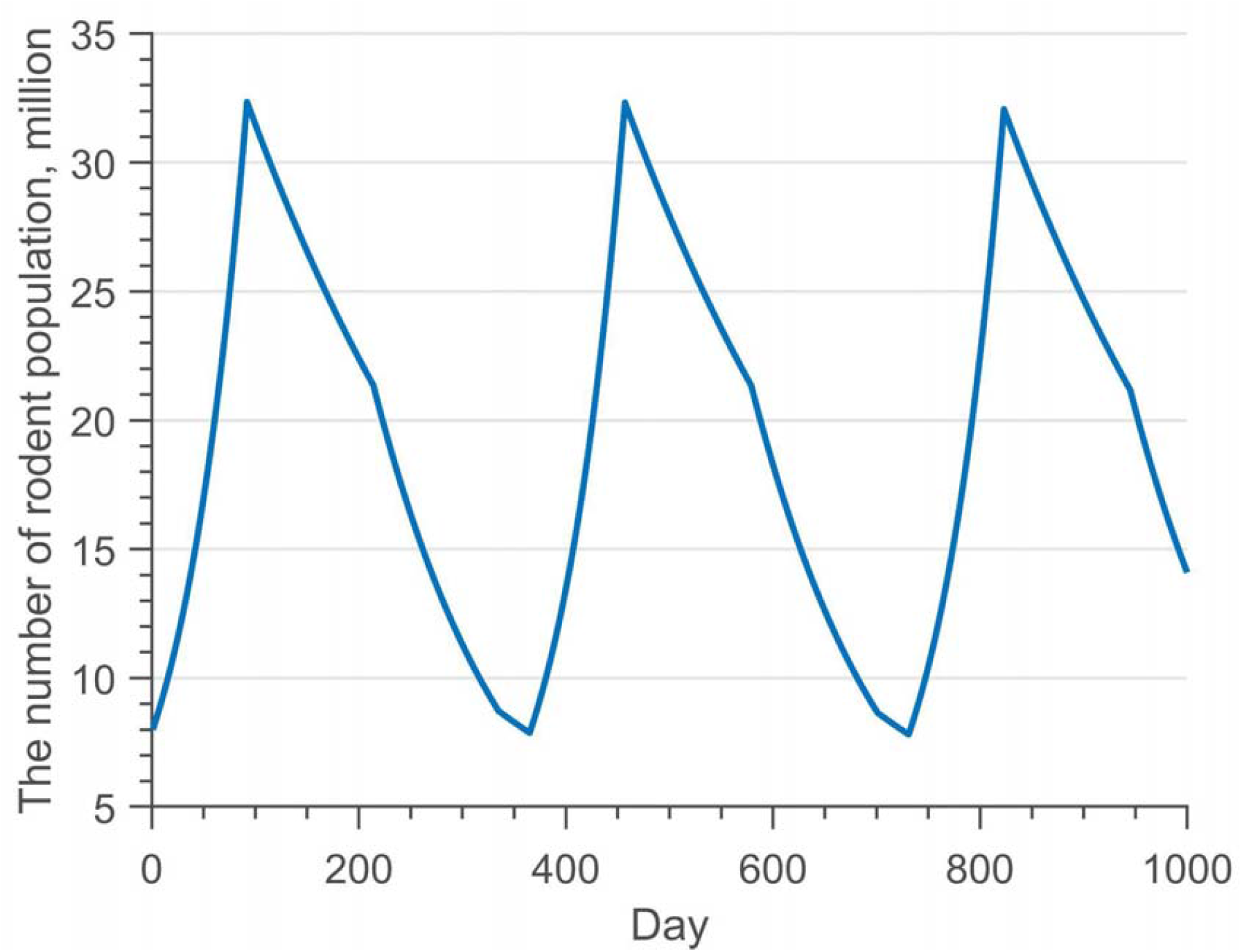
The seasonal dynamics of the rodent population. The simulation starts on May 1, 2022.

### Appendix D Sensitivity analysis

**Table A1:** Parameters range for the Latin Hypercube Sampling (LHS) in the sensitivity analysis

| Parameter |  | Definition | Range |
| --- | --- | --- | --- |
| Human related | $p_{HRG}$ | Proportion of the population that is in the HRG | [0.025, 0.05] |
| | $\beta_h$ | Probability of transmission per contact among HRG | [0.091, 0.214] |
| | $p_{c1}$ | Proportion of contact tracing in the LRG | [0.01, 0.6] |
| | $p_{c2}$ | Proportion of contact tracing in the HRG | [0.01, 1] |
| | $\theta_q$ | Proportion of infectious individuals with rash symptoms who seek medical help, be confirmed and isolated | [0.01, 0.8] |
| | $\alpha$ | Proportion of individuals who comply with the isolation strategy | [0.01, 0.5] |
| Rodent related | Nr | Total number of rodents population | [1000000, 10000000] |
| | $\beta_r$ | Product of contact rate among rodents and probability of transmission per contact among the rodent population | [0.091, 0.3] |
| | $\beta_{rh}$ | Product of contact rate between rodents and humans and probability of transmission per contact from the rodent to the human population | [0.00091, 0.003] |

